# Elevated risk of fibrotic hand manifestations in diabetes: a systematic review and meta-analysis of case-control studies

**DOI:** 10.1101/2025.04.21.25326143

**Authors:** Rucha Wagh, Kaustubh Chaudhari, Vanshaj Sharma, Sukeshini Khandagale, Rutvij Tope, Smita Dhadge, Stephanie Hendren, Maragatha Kuchibhatla, Chittaranjan Yajnik, Jennifer L. Ingram, Sanat Phatak

## Abstract

**Background:** Diabetes mellitus is associated with a cluster of hand manifestations—including Dupuytren’s disease (DD), flexor tenosynovitis (FT), limited joint mobility (LJM), and carpal tunnel syndrome (CTS)—collectively termed the “diabetic hand.” While these conditions share a common profibrotic histopathology, the magnitude and consistency of risk across populations remain unclear.

**Methods:** We reviewed and meta-analyzed case-control studies quantifying the risk of individual and combined fibrotic hand manifestations in individuals with diabetes compared to non-diabetic controls. We searched five databases (last updated August 2024) for case-control studies reporting prevalence of DD, FT, LJM, or CTS in individuals with diabetes. Pooled risk ratios (RRs) were calculated using a random-effects model. Subgroup analyses were conducted by diabetes type and publication year.

**Results:** Fifty-one case-control studies were included. In type 2 diabetes, pooled RRs were: DD 2.58 [1.85–3.60], FT 3.54 [2.07–6.05], LJM 3.18 [2.43–4.15], CTS 2.95 [1.83–4.77]. In type 1 diabetes, corresponding RRs were higher: DD 4.33 [1.69–11.12], FT 5.67 [2.25–14.30], LJM 3.98 [3.00–5.29], CTS 4.57 [2.46–8.49]. The combined risk of any hand fibrosis in type 2 diabetes was 3.39 [2.60–4.43]. Substantial heterogeneity was observed across studies.

**Conclusion:** Diabetes significantly increases the risk of all four fibrotic hand manifestations, with higher relative risks in type 1 diabetes. These findings probably support the concept of a shared fibrotic pathway and highlight the need for standardized diagnostic criteria and longitudinal assessment of musculoskeletal complications in diabetes.

**Protocol:** Registered prospectively with PROSPERO (CRD42022257285)

**Funding:** DBT/Wellcome India Alliance Fellowship (IA/CPHE/19/1/504607) to Dr Sanat Phatak

**Research in context:**

1. What is already known about this subject? Diabetes is associated with certain fibrotic hand conditions (Dupuytren’s disease (DD), flexor tenosynovitis (FT), limited joint mobility (LJM), and carpal tunnel syndrome (CTS)—collectively termed the diabetic hand). Hyperglycemia is known to be causal in developing these conditions but the relative risks and consistency across populations have not been systematically quantified.
2. What is the key question? What is the pooled risk of developing individual and combined hand manifestations in people with diabetes compared to non-diabetic controls?
3. What are the new findings?
  1. Diabetes increases the risk of Dupuytren’s disease, flexor tenosynovitis, limited joint mobility, and carpal tunnel syndrome. All four manifestations have similar relative risks.
  2. Relative risks were higher in patients with type 1 diabetes as compared to type 2 diabetes.
  3. Substantial heterogeneity in studies highlights the need for standardized definitions and quantification methods.
4. How might this impact on clinical practice in foreseeable future? Better recognition and measurement of the ‘diabetic hand’ as a clinical entity may prompt earlier diagnosis, functional assessment, and could serve as a screening tool for identifying a broader profibrotic tendency in diabetes.

## Introduction

Diabetes mellitus is linked with certain musculoskeletal manifestations in the hands that add a layer of morbidity to a chronic disease. Limited joint mobility (LJM), historically known as diabetic cheiroarthropathy, was initially identified in patients with type 1 diabetes. [1] Since then, a suite of manifestations including chronic flexor tenosynovitis (FTS), including the stenosing variety or trigger finger, carpal tunnel syndrome (CTS), and Dupuytren’s disease (DD) have been associated, collectively termed the “diabetic hand.” [2] All these conditions share a common histo-pathological feature, viz fibrotic proliferation in periarticular connective tissues in one of the flexor structures of the hand. Depending on the condition, excessive collagen deposition is seen in tendon sheaths (FTS), palmar fascia (DD), and skin (LJM). The prevalence of these hand complications in diabetes varies across studies.

Despite the growing recognition of these conditions, significant knowledge gaps remain. A recent Mendelian randomization study of data in the UK Biobank supports the role of hyperglycemia as a causal factor in these conditions. [3] Despite this finding, a marked variation in the condition exists even in long standing diabetes. On the flip side, these conditions are seen in prediabetes as well as individuals with normal glucose tolerance. [4] A lack of a unifying method of diagnosis and quantitation of hand fibrosis has hampered systematic evaluations of correlations with vascular and profibrotic conditions. Most studies have assessed the risk of individual manifestations, [5,6] and few report a combined risk of developing any individual condition. Finally, differences in risks in type 1 and type 2 diabetes may provide clues on causative factors and effect modifiers, considering different pathogeneses, inflammatory profiles, and life trajectories of these two broad subtypes.

A meta-analysis can inform the magnitude and consistency of risk across populations. We aimed to address some of these gaps by synthesizing data from case-control studies, to quantify the risk for each manifestation as well as that for combined hand involvement. We quantified the risk of key hand manifestations in individuals with diabetes compared to non-diabetic controls in the different subtypes of diabetes, and reported pooled risk ratios individually for all four hand manifestations.

## Methods

### Search strategy

A comprehensive search strategy was developed to identify relevant studies on the topic of hand manifestations in diabetes mellitus (ESM Figure 1). The first search was conducted on 7 August 2021 using the following electronic databases: Medline (PubMed), Embase (Elsevier), CINAHL (EBSCOhost), Scopus (Elsevier), and Clinicaltrials.gov. A follow up search was conducted on 1 August 2024.

**ESM Figure 1:** PRISMA flow diagram for study selection process

The search terms included a combination of keywords and Medical Subject Headings related to prevalence of hand manifestation in diabetes mellitus, a detailed search strategy has been mentioned in ESM Table 2. We included original articles presenting the prevalence of 4 outcomes (LJM, DD, FTS, CTS) in diabetes mellitus (all types) written in the English language. We excluded studies having patients with other systemic fibrotic illnesses (scleroderma, nephrogenic systemic fibrosis) and other diseases affecting the hand (rheumatoid arthritis, psoriatic arthritis). Two independent reviewers (KC, physician and RW, biology researcher) performed screening, and a third independent reviewer (SP, physician) was assigned for consensus decision.

For the excluded review articles, we performed a separate search of the references cited in these articles to account for any new article which was missed in our initial search. Since three years had elapsed after the initial literature search was performed, we updated the search for the period of August 2021 to August 2024 using the same search strategy. A detailed overview of the search flow and the PRISM checklist have been provided in the ESM Figure 1 and ESM Table 1, respectively.

### Data extraction

Two independent researchers were involved in the data extraction, and a third independent reviewer was assigned for consensus decision. The final data sheet considered for data cleaning and analysis was of the consensus.

### Statistical analysis

We aimed to estimate the probability of developing individual hand manifestations in patients with diabetes compared to those without diabetes. This probability, expressed as a risk ratio (RR), represents the likelihood of a hand manifestation occurring in individuals with diabetes relative to those without diabetes. To ensure methodological consistency, we restricted our analysis to case-control studies, selecting these from the broader study set.

We conducted a meta-analysis using a case-control strategy. RRs were calculated for each outcome and study, quantifying the relative risk of hand manifestations in participants with diabetes compared to those without diabetes. A random-effects model was used to pool the RRs and compute 95% confidence intervals (CIs), accounting for between-study variability. Between-study heterogeneity was incorporated using the Restricted Maximum Likelihood Estimation (REML) for variance (τ²). Study-specific risk ratios and their variances were log-transformed for analysis, and pooled estimates were obtained using inverse-variance weighting, where studies with lower variance were assigned greater weight. The pooled RRs were back-transformed for interpretation.

Forest plots were generated to visualize individual study estimates and the overall pooled effect, along with heterogeneity measures such as Cochrane’s Q-statistic and I². We considered an I² value greater than 40% as indicative of considerable heterogeneity. [7] The Z-score was calculated to assess the significance of the test statistics for the risk ratios, with positive Z-values indicating the strength of association. A p-value <0.05 was considered statistically significant, indicating the need for further bias assessment if heterogeneity was substantial.

We used Covidence Systematic Review Software (Veritas Health Innovation, Melbourne, Australia; Available at www.covidence.org) for data management, including search and data extraction. All statistical analyses were performed in R software using the ‘Meta’ package.

### Heterogeneity Assessment and risk of publication bias

Qualitative assessment was performed using the Newcastle-Ottawa scale (NOS) for case-control study design. Each study was scored based on eight criteria and a composite score was also mentioned. The eight scores were based on three domains: selection of study groups (four stars), comparability of study groups (two stars), and ascertainment of exposures and outcomes (three stars); thus, a maximum of nine stars could be obtained for each case-control study. A threshold score to differentiate good and poor-quality studies has not been developed yet and hence the NOS can be used to compare the quality of studies across domains and in total.

#### Ethics

This study has received ethics approval from the KEM Hospital Research Center Ethics Committee (KEMHRC/RVC/EC/1518) and is registered prospectively with PROSPERO (CRD42022257285)

## Results

Following the initial search across the 5 databases, a total of 12361 records were identified. After removing duplicates, 7233 unique records were screened for eligibility based on their titles and abstracts (ESM Figure 1). From the extracted dataset, 51 studies were of case-control design which had provided the prevalence for DD, FT, LJM, CTS (at least one) and could be statistically analysed. Of 51, 28 studies reported data on type 2 diabetes, 23 studies reported type 1 diabetes, and seven did not specify type of diabetes mellitus (Table 1).

**Table 1:**
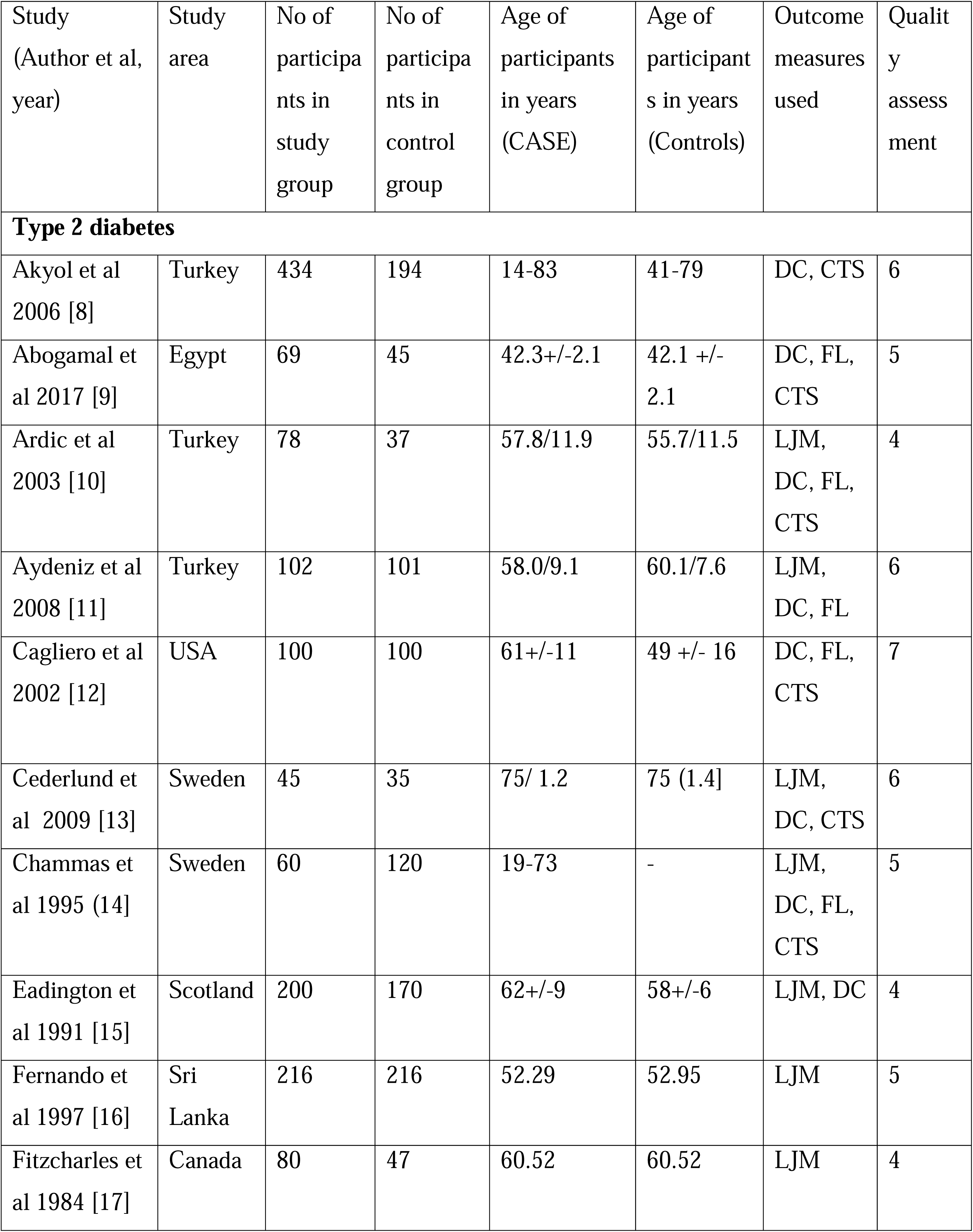
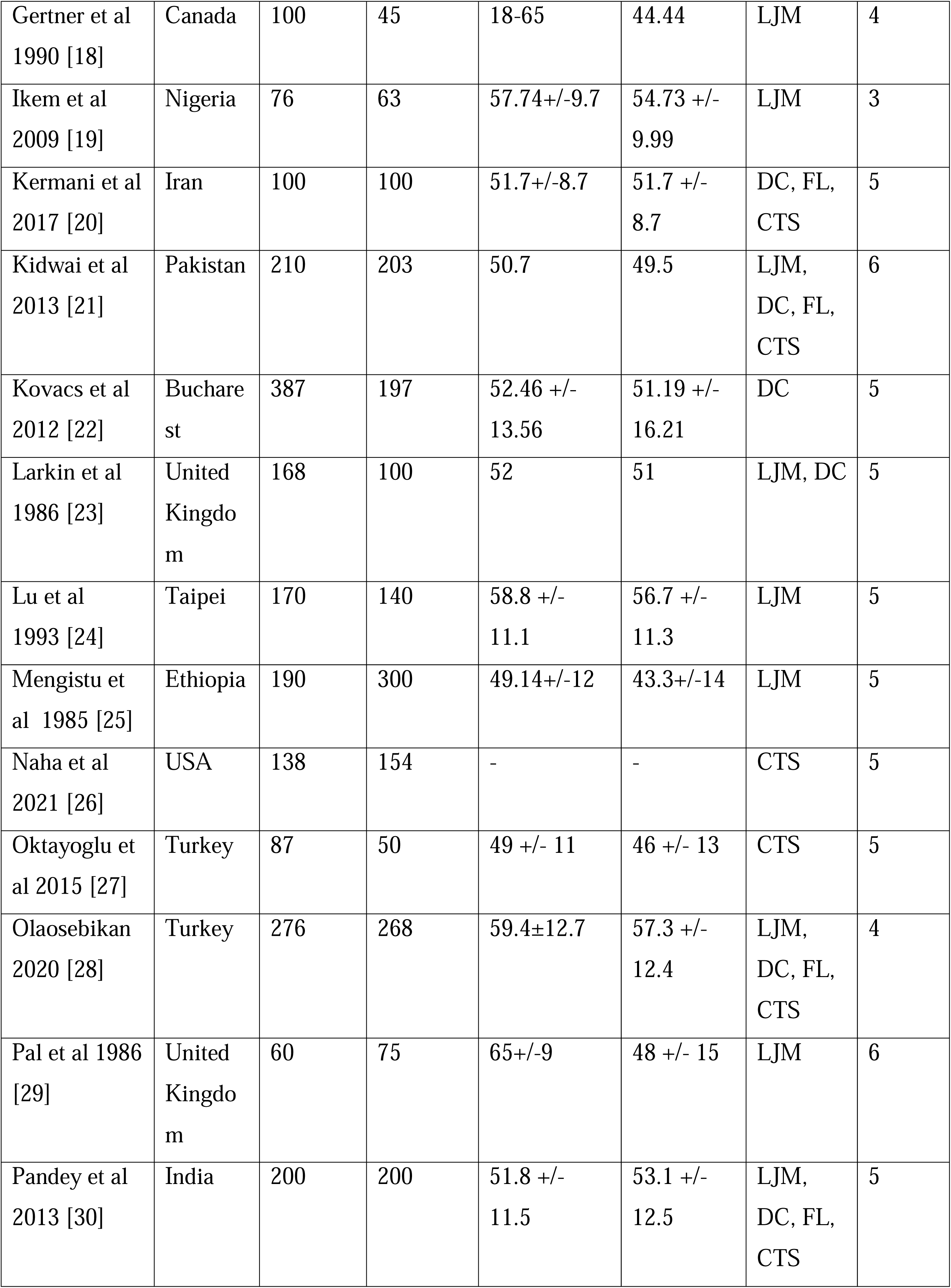
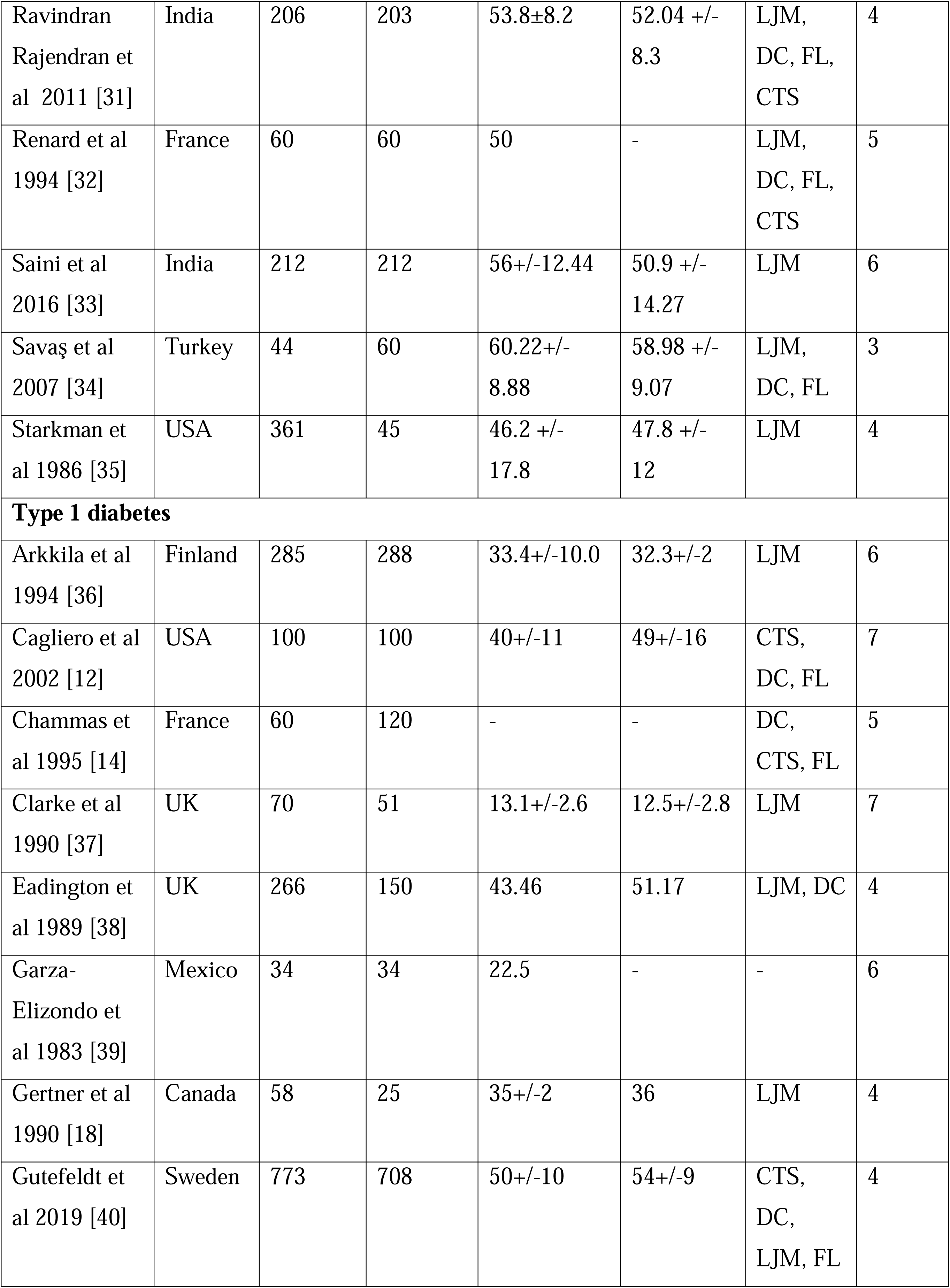
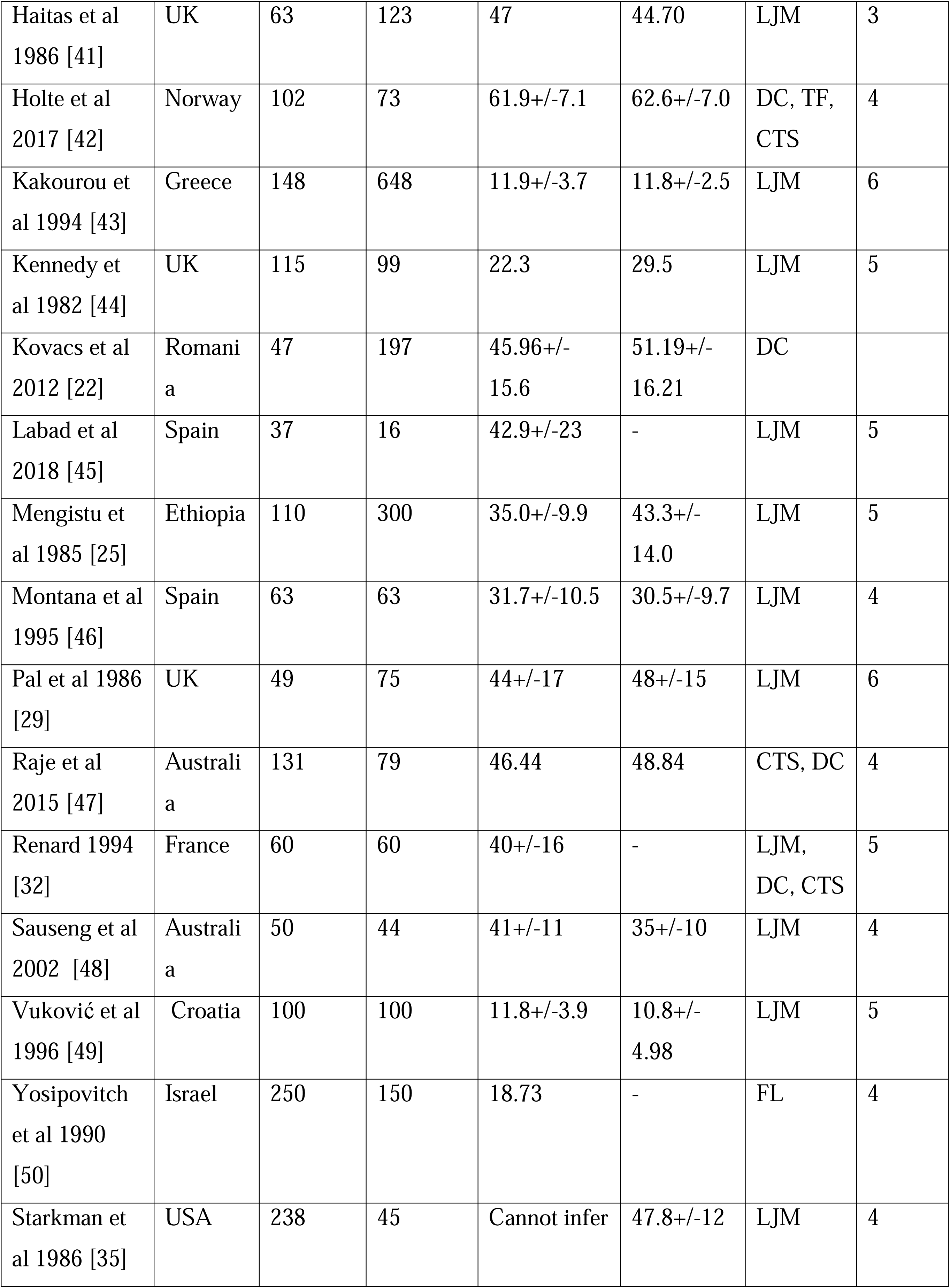
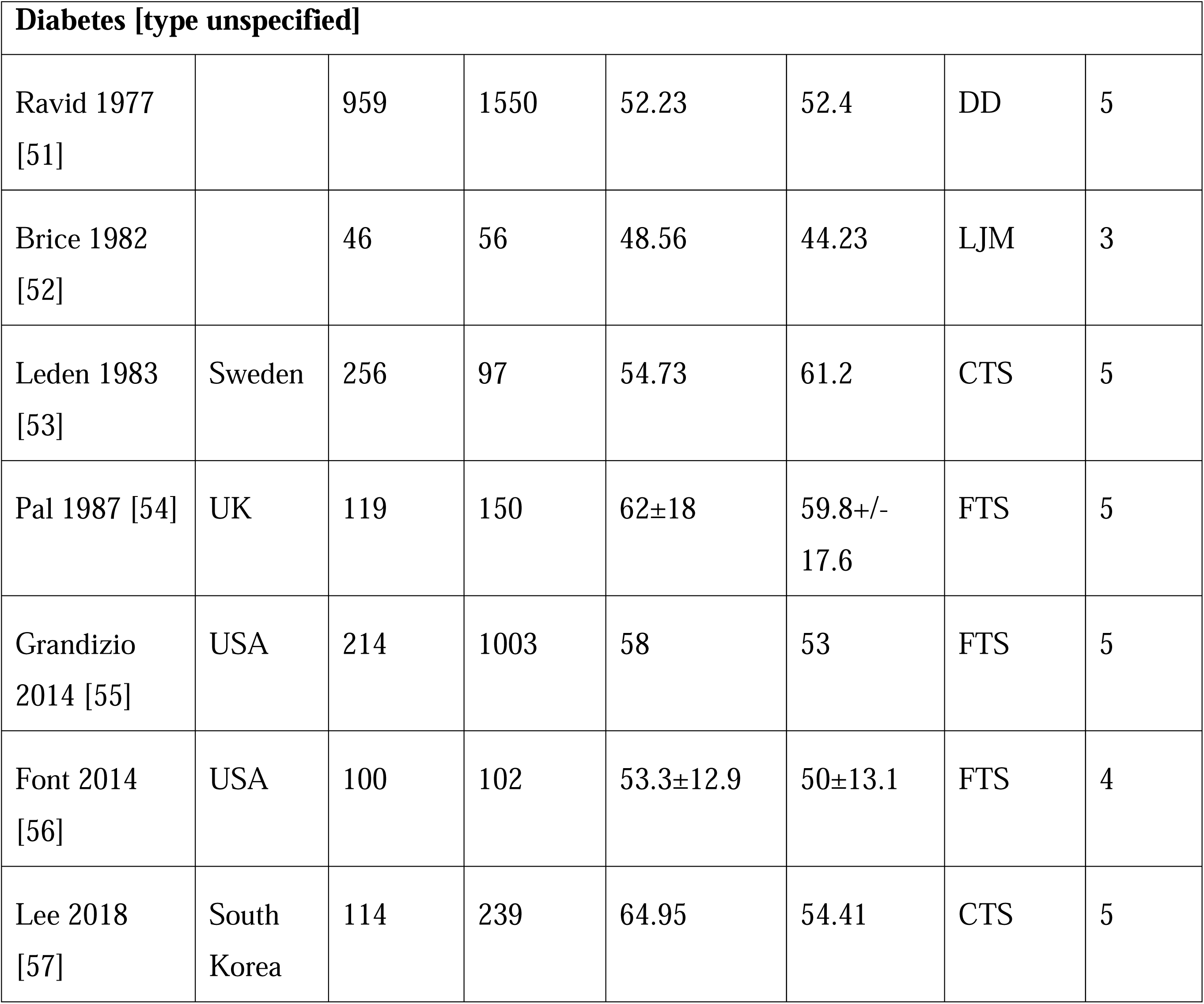
Characteristics of the studies included in the review.

| Study<br>(Author et al,<br>year) | Study<br>area | No of<br>participa<br>nts in<br>study<br>group | No of<br>participa<br>nts in<br>control<br>group | Age of<br>participants<br>in years<br>(CASE) | Age of<br>participant<br>s in years<br>(Controls) | Outcome<br>measures<br>used | Qualit<br>y<br>assess<br>ment |
| --- | --- | --- | --- | --- | --- | --- | --- |
| **Type 2 diabetes** |  |  |  |  |  |  |  |
| Akyol et al<br>2006 [8] | Turkey | 434 | 194 | 14-83 | 41-79 | DC, CTS | 6 |
| Abogamal et<br>al 2017 [9] | Egypt | 69 | 45 | 42.3+/-2.1 | 42.1 +/-<br>2.1 | DC, FL,<br>CTS | 5 |
| Ardic et al<br>2003 [10] | Turkey | 78 | 37 | 57.8/11.9 | 55.7/11.5 | LJM,<br>DC, FL,<br>CTS | 4 |
| Aydeniz et al<br>2008 [11] | Turkey | 102 | 101 | 58.0/9.1 | 60.1/7.6 | LJM,<br>DC, FL | 6 |
| Cagliero et al<br>2002 [12] | USA | 100 | 100 | 61+/-11 | 49 +/- 16 | DC, FL,<br>CTS | 7 |
| Cederlund et<br>al 2009 [13] | Sweden | 45 | 35 | 75/ 1.2 | 75 (1.4] | LJM,<br>DC, CTS | 6 |
| Chammas et<br>al 1995 (14] | Sweden | 60 | 120 | 19-73 | - | LJM,<br>DC, FL,<br>CTS | 5 |
| Eadington et<br>al 1991 [15] | Scotland | 200 | 170 | 62+/-9 | 58+/-6 | LJM, DC | 4 |
| Fernando et<br>al 1997 [16] | Sri<br>Lanka | 216 | 216 | 52.29 | 52.95 | LJM | 5 |
| Fitzcharles et<br>al 1984 [17] | Canada | 80 | 47 | 60.52 | 60.52 | LJM | 4 |
| Gertner et al<br>1990 [18] | Canada | 100 | 45 | 18-65 | 44.44 | LJM | 4 |
| Ikem et al<br>2009 [19] | Nigeria | 76 | 63 | 57.74+/-9.7 | 54.73 +/-<br>9.99 | LJM | 3 |
| Kermani et al<br>2017 [20] | Iran | 100 | 100 | 51.7+/-8.7 | 51.7 +/-<br>8.7 | DC, FL,<br>CTS | 5 |
| Kidwai et al<br>2013 [21] | Pakistan | 210 | 203 | 50.7 | 49.5 | LJM,<br>DC, FL,<br>CTS | 6 |
| Kovacs et al<br>2012 [22] | Bucharest | 387 | 197 | 52.46 +/-<br>13.56 | 51.19 +/-<br>16.21 | DC | 5 |
| Larkin et al<br>1986 [23] | United Kingdom | 168 | 100 | 52 | 51 | LJM, DC | 5 |
| Lu et al<br>1993 [24] | Taipei | 170 | 140 | 58.8 +/-<br>11.1 | 56.7 +/-<br>11.3 | LJM | 5 |
| Mengistu et al<br>1985 [25] | Ethiopia | 190 | 300 | 49.14+/-12 | 43.3+/-14 | LJM | 5 |
| Naha et al<br>2021 [26] | USA | 138 | 154 | - | - | CTS | 5 |
| Oktayoglu et al<br>2015 [27] | Turkey | 87 | 50 | 49 +/- 11 | 46 +/- 13 | CTS | 5 |
| Olaosebikan<br>2020 [28] | Turkey | 276 | 268 | 59.4±12.7 | 57.3 +/-<br>12.4 | LJM,<br>DC, FL,<br>CTS | 4 |
| Pal et al 1986<br>[29] | United Kingdom | 60 | 75 | 65+/-9 | 48 +/- 15 | LJM | 6 |
| Pandey et al<br>2013 [30] | India | 200 | 200 | 51.8 +/-<br>11.5 | 53.1 +/-<br>12.5 | LJM,<br>DC, FL,<br>CTS | 5 |
| Ravindran<br>Rajendran et<br>al 2011 [31] | India | 206 | 203 | 53.8±8.2 | 52.04 +/-<br>8.3 | LJM,<br>DC, FL,<br>CTS | 4 |
| Renard et al<br>1994 [32] | France | 60 | 60 | 50 | - | LJM,<br>DC, FL,<br>CTS | 5 |
| Saini et al<br>2016 [33] | India | 212 | 212 | 56+/-12.44 | 50.9 +/-<br>14.27 | LJM | 6 |
| Savaş et al<br>2007 [34] | Turkey | 44 | 60 | 60.22+/-<br>8.88 | 58.98 +/-<br>9.07 | LJM,<br>DC, FL | 3 |
| Starkman et<br>al 1986 [35] | USA | 361 | 45 | 46.2 +/-<br>17.8 | 47.8 +/-<br>12 | LJM | 4 |
| **Type 1 diabetes** |  |  |  |  |  |  |  |
| Arkkila et al<br>1994 [36] | Finland | 285 | 288 | 33.4+/-10.0 | 32.3+/-2 | LJM | 6 |
| Cagliero et al<br>2002 [12] | USA | 100 | 100 | 40+/-11 | 49+/-16 | CTS,<br>DC, FL | 7 |
| Chammas et<br>al 1995 [14] | France | 60 | 120 | - | - | DC,<br>CTS, FL | 5 |
| Clarke et al<br>1990 [37] | UK | 70 | 51 | 13.1+/-2.6 | 12.5+/-2.8 | LJM | 7 |
| Eadington et<br>al 1989 [38] | UK | 266 | 150 | 43.46 | 51.17 | LJM, DC | 4 |
| Garza-Elizondo et<br>al 1983 [39] | Mexico | 34 | 34 | 22.5 | - | - | 6 |
| Gertner et al<br>1990 [18] | Canada | 58 | 25 | 35+/-2 | 36 | LJM | 4 |
| Gutefeldt et<br>al 2019 [40] | Sweden | 773 | 708 | 50+/-10 | 54+/-9 | CTS,<br>DC,<br>LJM, FL | 4 |
| Haitas et al<br>1986 [41] | UK | 63 | 123 | 47 | 44.70 | LJM | 3 |
| Holte et al<br>2017 [42] | Norway | 102 | 73 | 61.9+/-7.1 | 62.6+/-7.0 | DC, TF,<br>CTS | 4 |
| Kakourou et<br>al 1994 [43] | Greece | 148 | 648 | 11.9+/-3.7 | 11.8+/-2.5 | LJM | 6 |
| Kennedy et<br>al 1982 [44] | UK | 115 | 99 | 22.3 | 29.5 | LJM | 5 |
| Kovacs et al<br>2012 [22] | Romani<br>a | 47 | 197 | 45.96+/-<br>15.6 | 51.19+/-<br>16.21 | DC |  |
| Labad et al<br>2018 [45] | Spain | 37 | 16 | 42.9+/-23 | - | LJM | 5 |
| Mengistu et<br>al 1985 [25] | Ethiopia | 110 | 300 | 35.0+/-9.9 | 43.3+/-<br>14.0 | LJM | 5 |
| Montana et al<br>1995 [46] | Spain | 63 | 63 | 31.7+/-10.5 | 30.5+/-9.7 | LJM | 4 |
| Pal et al 1986<br>[29] | UK | 49 | 75 | 44+/-17 | 48+/-15 | LJM | 6 |
| Raje et al<br>2015 [47] | Australi<br>a | 131 | 79 | 46.44 | 48.84 | CTS, DC | 4 |
| Renard 1994<br>[32] | France | 60 | 60 | 40+/-16 | - | LJM,<br>DC, CTS | 5 |
| Sauseng et al<br>2002 [48] | Australi<br>a | 50 | 44 | 41+/-11 | 35+/-10 | LJM | 4 |
| Vuković et al<br>1996 [49] | Croatia | 100 | 100 | 11.8+/-3.9 | 10.8+/-<br>4.98 | LJM | 5 |
| Yosipovitch<br>et al 1990<br>[50] | Israel | 250 | 150 | 18.73 | - | FL | 4 |
| Starkman et<br>al 1986 [35] | USA | 238 | 45 | Cannot infer | 47.8+/-12 | LJM | 4 |

| **Diabetes [type unspecified]** |  |  |  |  |  |  |  |
| --- | --- | --- | --- | --- | --- | --- | --- |
| Ravid 1977<br>[51] |  | 959 | 1550 | 52.23 | 52.4 | DD | 5 |
| Brice 1982<br>[52] |  | 46 | 56 | 48.56 | 44.23 | LJM | 3 |
| Leden 1983<br>[53] | Sweden | 256 | 97 | 54.73 | 61.2 | CTS | 5 |
| Pal 1987 [54] | UK | 119 | 150 | 62±18 | 59.8+/-<br>17.6 | FTS | 5 |
| Grandizio<br>2014 [55] | USA | 214 | 1003 | 58 | 53 | FTS | 5 |
| Font 2014<br>[56] | USA | 100 | 102 | 53.3±12.9 | 50±13.1 | FTS | 4 |
| Lee 2018<br>[57] | South<br>Korea | 114 | 239 | 64.95 | 54.41 | CTS | 5 |

### Risk of hand manifestations in Type 2 diabetes

Out of the 28 studies, prevalence for DD, FT, LJM and CTS in both groups were provided in 13, 10, 18 and 12 studies, respectively. All the studies which had 0 prevalence in either group were excluded from the analysis.

The pooled risk ratio with the corresponding CI for DD, FT, LJM, CTS was 2.58 [1.85; 3.60], 3.54 [2.07; 6.05], 3.18 [2.43; 4.15], 2.95 [1.83; 4.77], respectively. A significant level of heterogeneity was observed during the meta-analysis for DD (I2=63.5%, p<0.01), LJM (I2=77.8%, p<0.01), CTS (I2=63.9%, p<0.01), respectively, but not for FT (I2=29.1%, p=0.177) (Figure 1).

**Figure 1:**
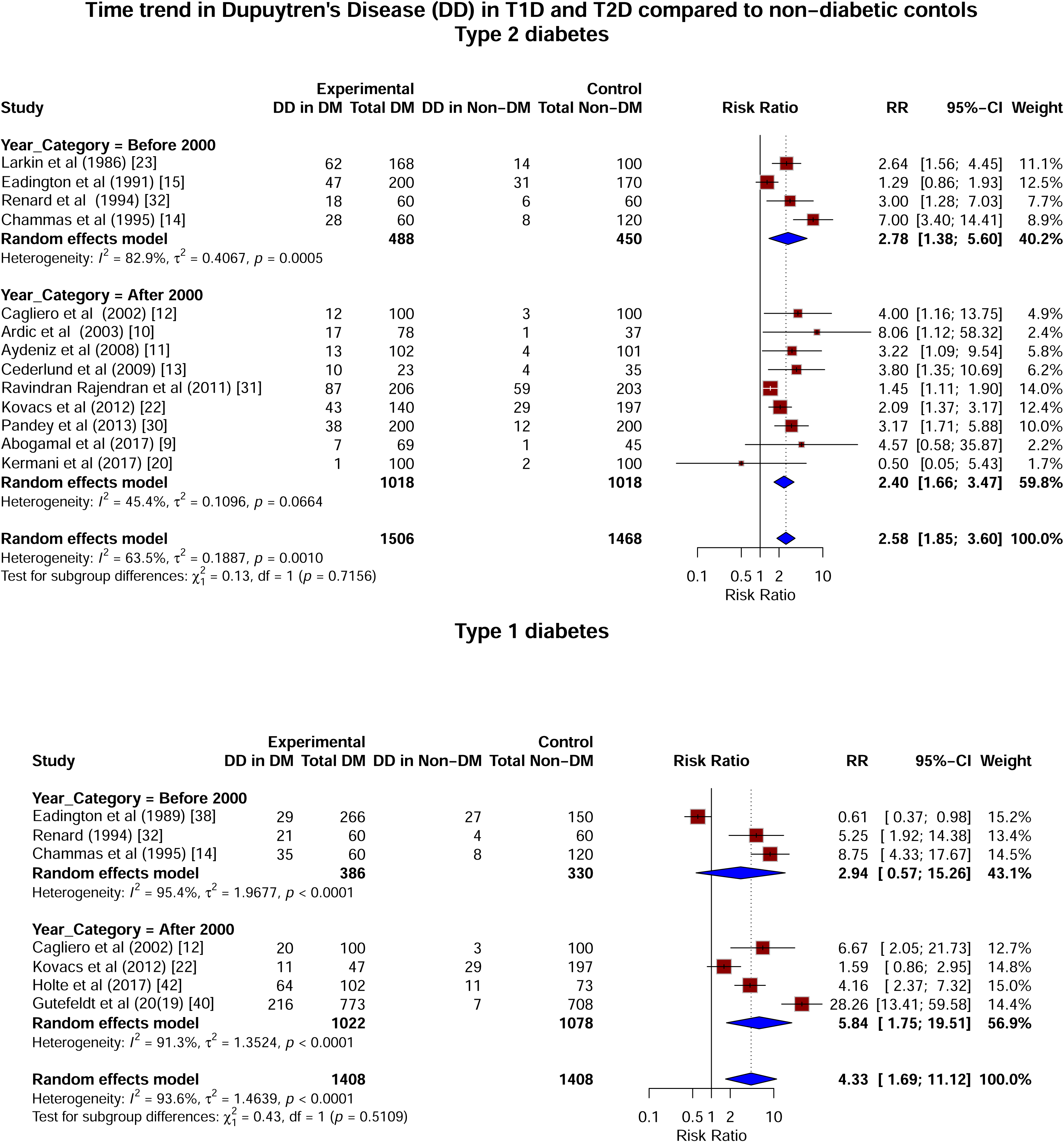
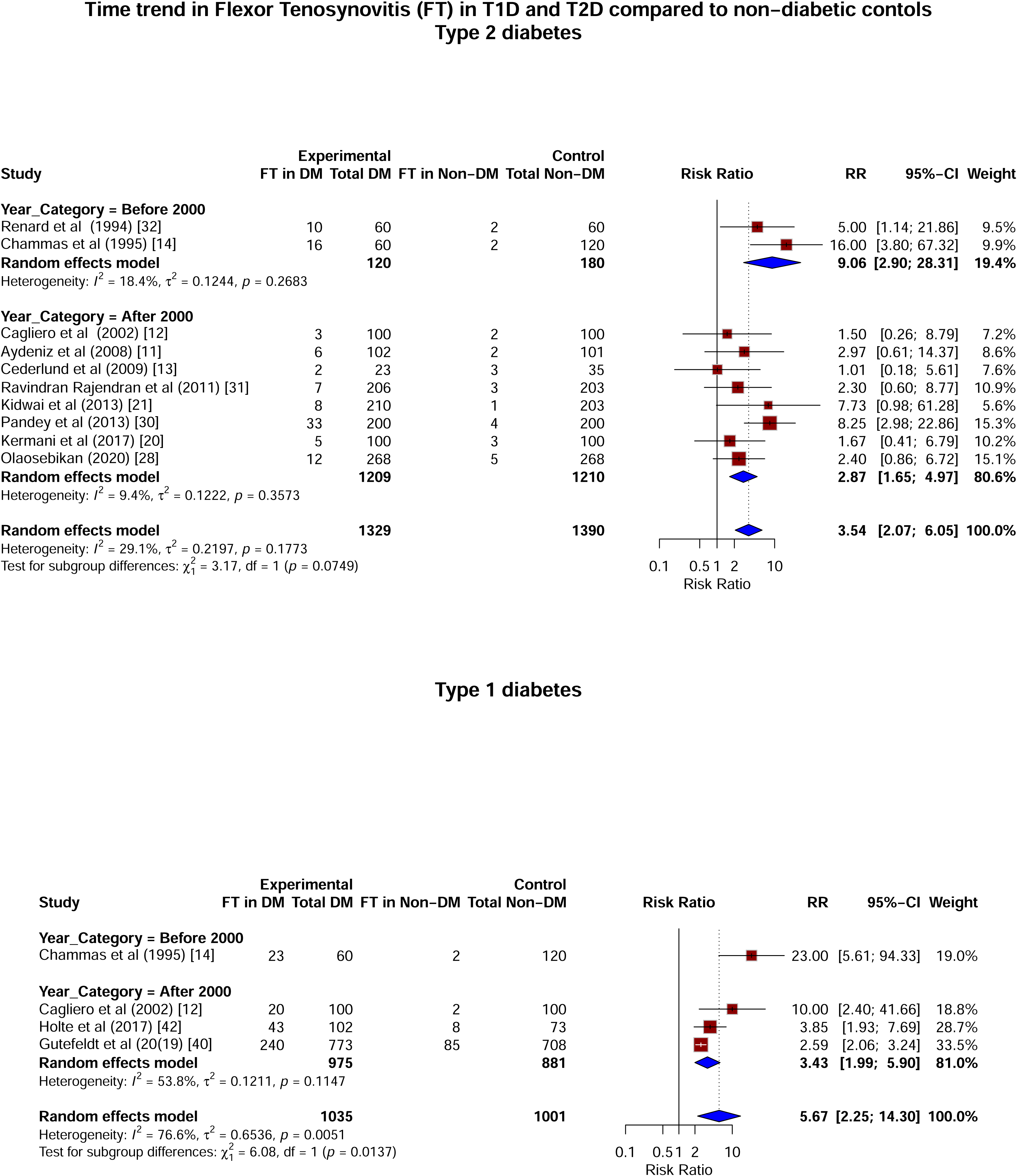
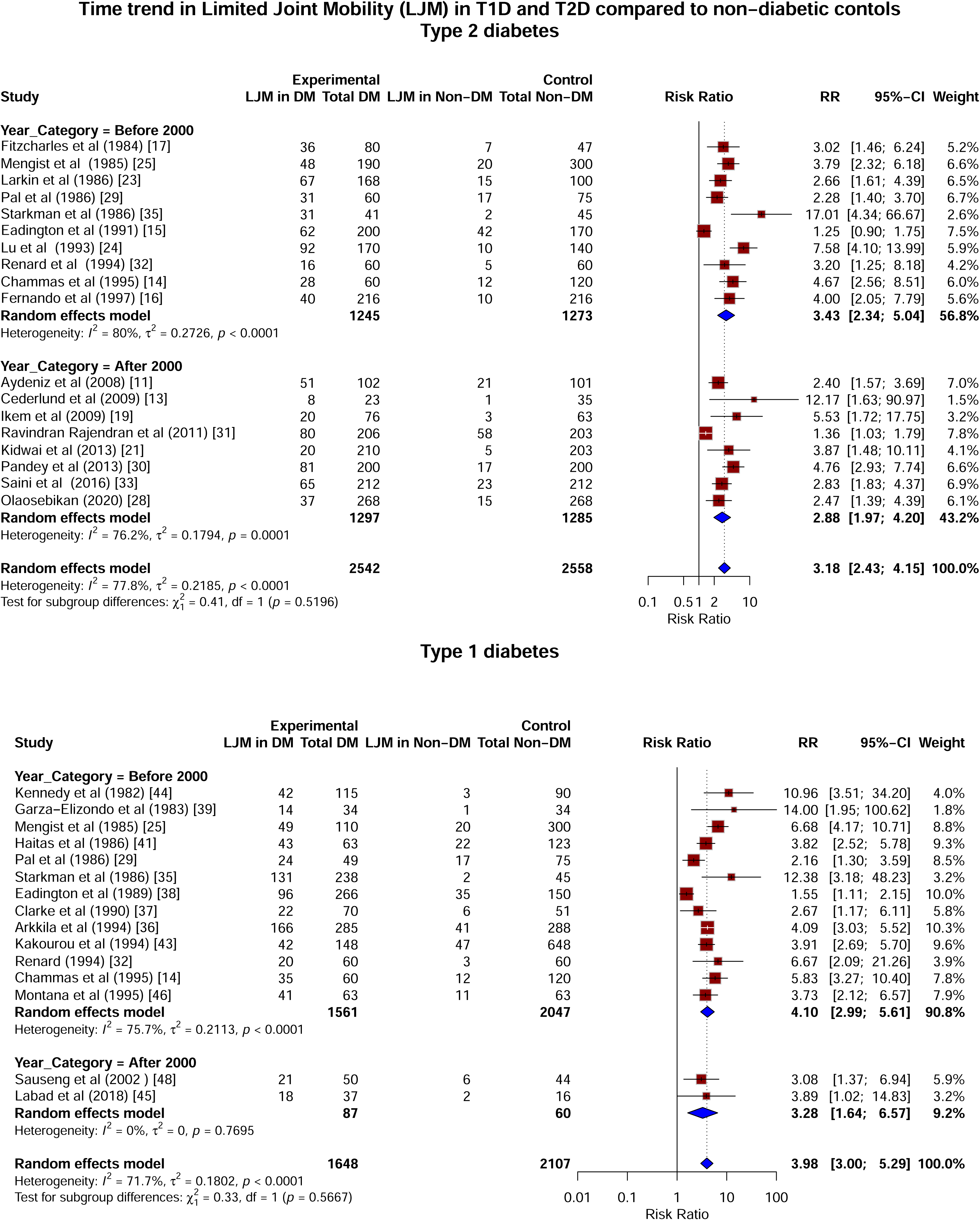
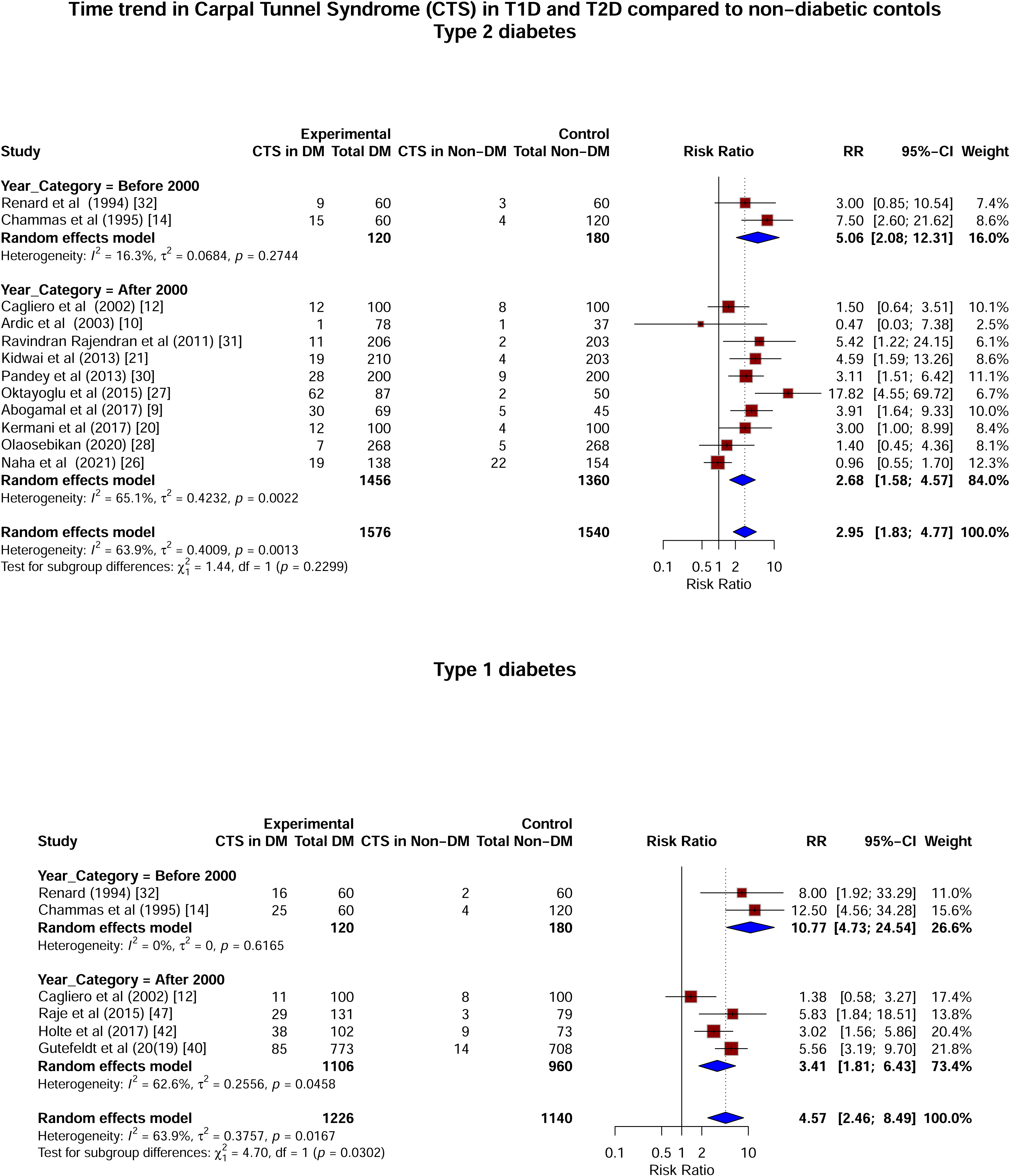
Forest Plots for each hand outcome and type of diabetes: Forest plots presenting risk ratios of the hand manifestations in participants with diabetes compared to participants without diabetes separately for type 2 diabetes and type 1 diabetes. (A) Dupuytren’s in diabetes vs non-diabetes, (B) Flexor tenosynovitis in diabetes vs non-diabetes, (C) Limited joint mobility in diabetes vs non-diabetes, (D) Carpal tunnel syndrome in diabetes vs non-diabetes

**Figure 2:**
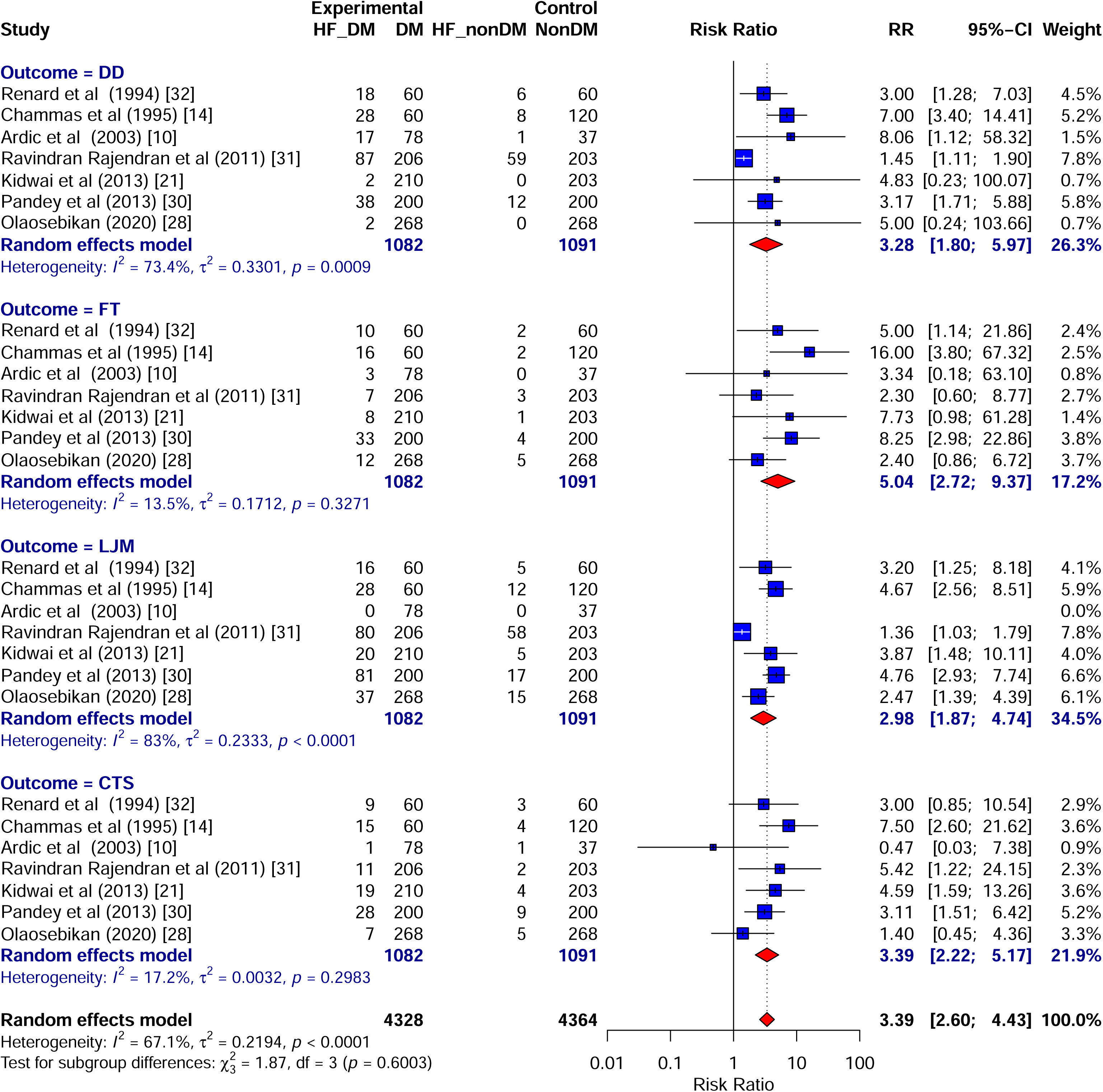
Forest plot for combined prevalence of 4 outcomes in type 2 diabetes compared to non-diabetic controls

Seven studies provided prevalence for all the hand outcomes in both the groups. Hence, we examined for the cumulative risk of these outcomes grouped as ‘Hand fibrosis (HF)’, which was 3.39 [2.60; 4.43] compared to controls with a significant heterogeneity of 67.1% (p<0.01).

### Risk of hand manifestations in Type 1 diabetes

Out of the 23 studies, prevalence for DD, FT, LJM and CTS in both groups were provided in 7, 4, 15 and 6 studies, respectively. Studies showing 0 prevalence for either cases or controls were excluded.

The pooled risk ratio with their corresponding CI for DD, FT, LJM, CTS were 4.33 [1.69; 11.12], 5.67 [2.25; 14.30], 3.98 [3.00; 5.29], 4.57 [2.46; 8.49], respectively. However, a significant level of heterogeneity exists during the meta-analysis for DD (I^2^=93.6%, p value<0.01), FT (I^2^=76.6%, p value<0.01), LJM (I^2^=71.7%, p value<0.01), CTS (I^2^=63.9%, p=0.016), respectively (Figure 1).

Only 2 studies provided prevalence for all the hand outcomes in both groups. Given the small number, we could not assess the cumulative risk of all outcomes together in type 1 diabetes.

### Risk ratios according to year of publication

In a secondary analysis, we also checked if there was any trend in the RR across the years. It showed that for type 2 diabetes the RRs were higher before the year 2000, which fell consistently for all the 4 outcomes post-2000 (Table 2, Figure 3). For type 1 diabetes, no consistent trend was observed before and after the year 2000, potentially due to small numbers of studies.

**Figure 3:**
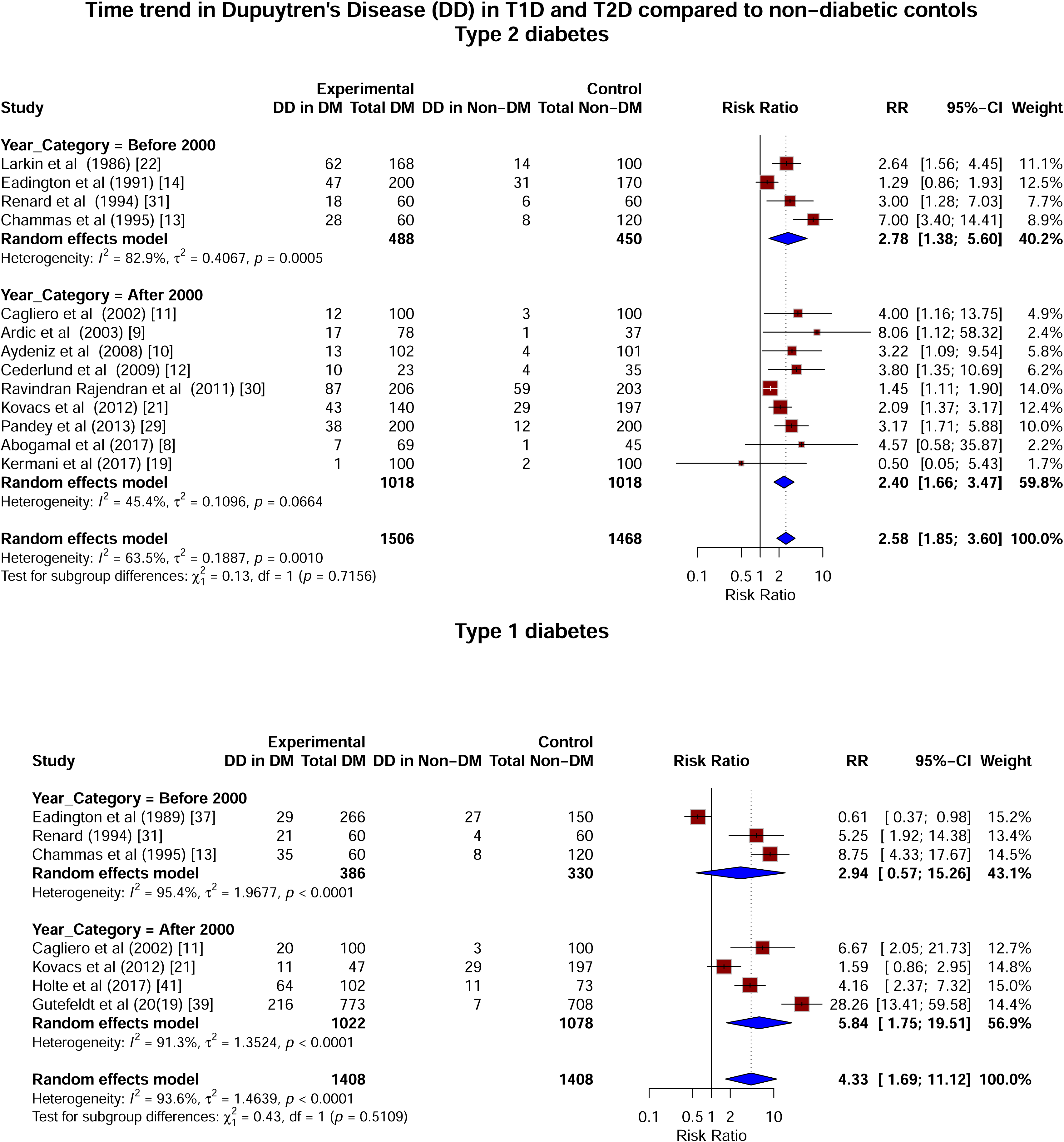
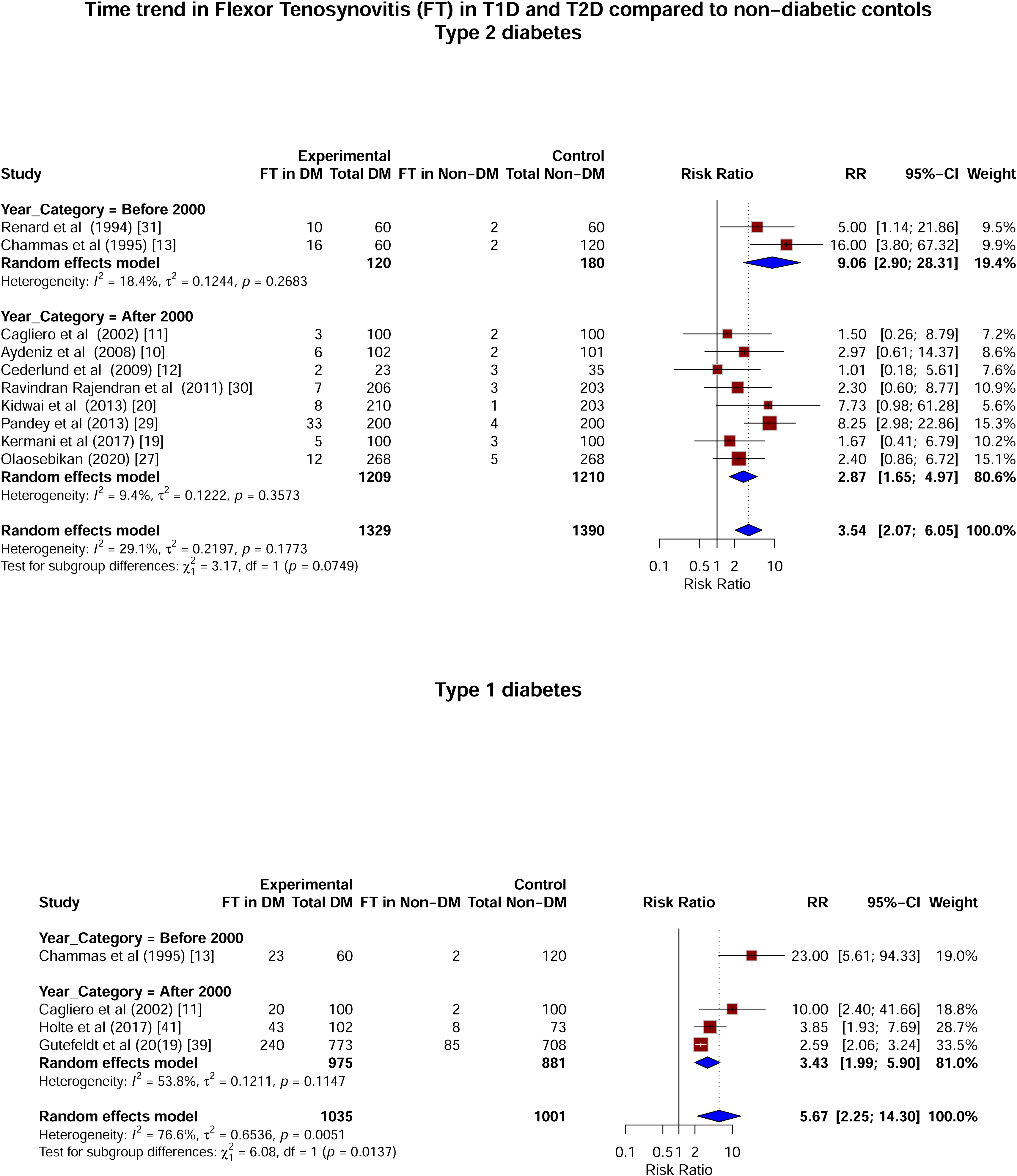
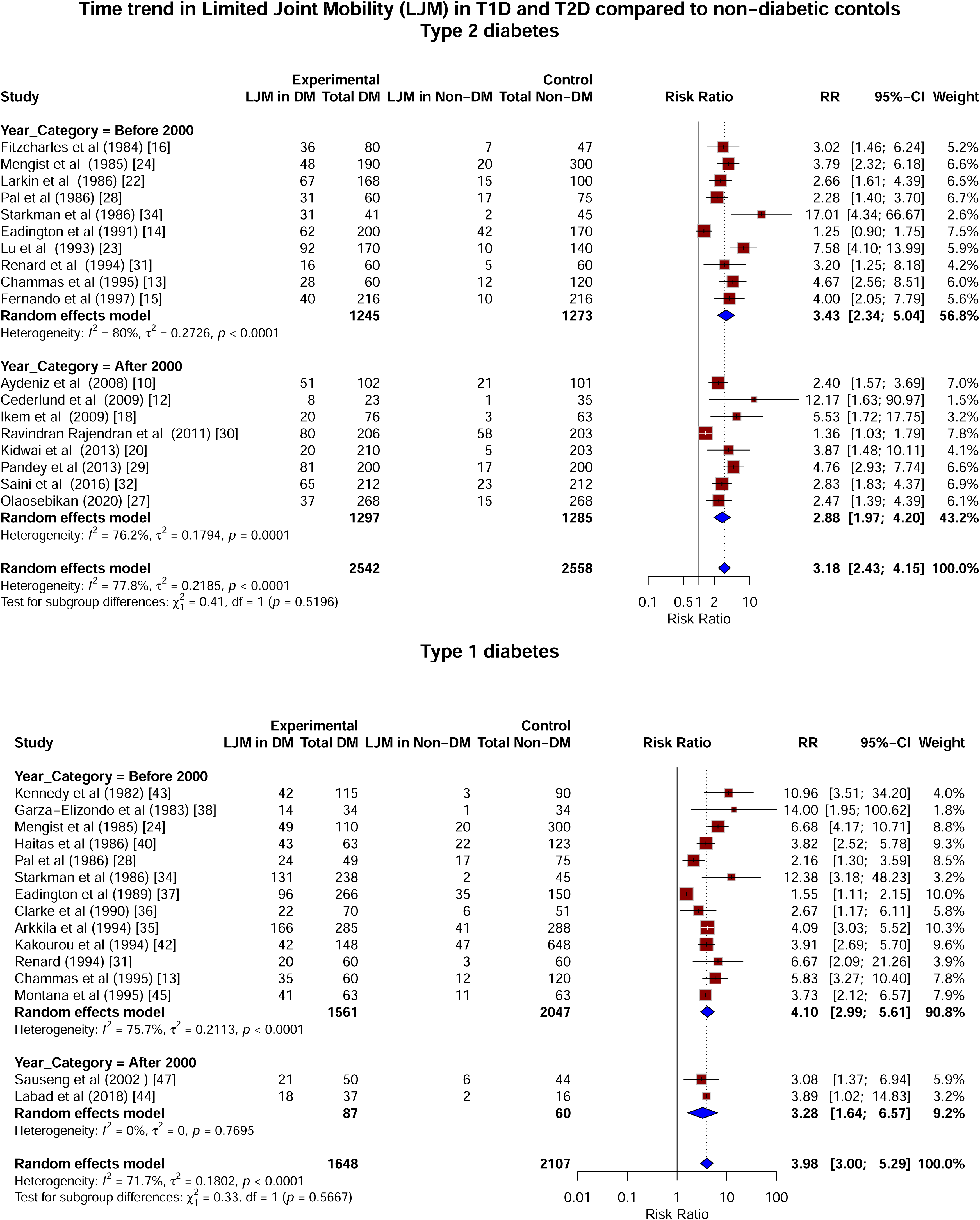
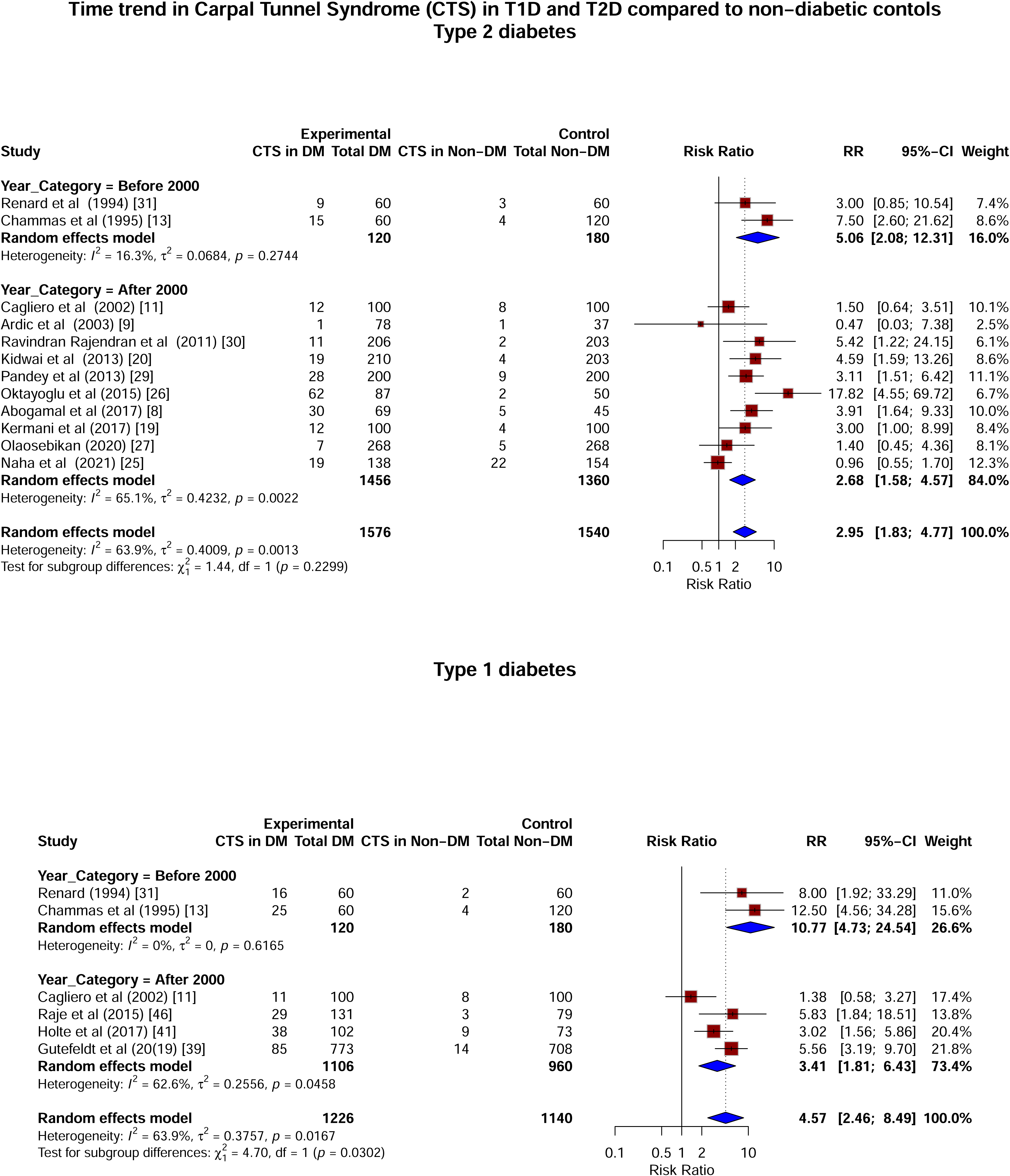
Forest plots of time trends of hand manifestations in diabetes versus controls (before and after 2000) (A) Dupuytren’s in diabetes vs non-diabetes, (B) Flexor tenosynovitis in diabetes vs non-diabetes, (C) Limited joint mobility in diabetes vs non-diabetes, (D) Carpal tunnel syndrome in diabetes vs non-diabetes

**Table 2:** Trend of risk ratios according to years (before and after 2000)

| Type | DD | FT | LJM | CTS |
| --- | --- | --- | --- | --- |
| Type 1 diabetes |  |  |  |  |
| Before year 2000 | 2.94 [0.57, 15.26] | - | 4.10 [2.99; 5.61] | 10.77 [4.73, 24.54] |
| After year 2000 | 5.84 [1.75, 19.51] | 3.43 [1.99; 5.90] | 3.28 [1.64; 6.57] | 3.41 [1.81; 6.43] |
| Type 2 diabetes |  |  |  |  |
| Before year 2000 | 2.78 [1.38, 5.60] | 9.06 [2.90; 28.31] | 3.43 [2.34; 5.04] | 5.06 [2.08; 12.31] |
| After year 2000 | 2.40 [1.66, 3.47] | 2.87 [1.65; 4.97] | 2.88 [1.97; 4.20] | 2.68 [1.58; 4.57] |

### Diabetes (type unspecified)

Of 7 studies which did not specify the diabetes type, prevalence for DD, FT, LJM and CTS was mentioned in 1, 3, 2, and 1, respectively. Given the small numbers, we could not perform meta-analysis for this group.

### Qualitative Assessment

The qualitative assessment results are presented in both composite and individual score format (ESM Table 3). Most studies (22 out of 23 studies on type 1 diabetes, 26 out of 28 studies on type 2 diabetes, and 6 out of 7 studies on unspecified diabetes) received four or more stars. Overall, 26 out of 51 studies have provided clear definitions for cases and only 3 studies have commented on how non-respondents were identified.

## Discussion

In this systematic review of case-control studies, we confirmed that diabetes increased the risk of developing each one of the four described phenotypes of the ‘diabetic hand’, namely LJM, DD, FT, and CTS. Using pooled risk ratios across studies, we found that the risk of all these manifestations was increased about three-fold in type 2 diabetes, and between four- and five-fold in type 1 diabetes. Cumulatively, the risk of developing any hand manifestation in type 2 diabetes was more than three-fold. Substantial heterogeneity in the studies was observed.

Fibrosis in diabetes is not restricted to the hands but has been observed in multiple organs, including the kidneys, liver, and lungs. Individuals with diabetes have a significantly increased risk of liver fibrosis, (adjusted odds ratio of 3.02 compared to non-diabetic individuals). [58] Similarly, diabetes has been implicated as a risk factor for idiopathic pulmonary fibrosis, with studies reporting higher mortality in affected individuals (hazard ratio 2.5, 95% CI 1.04–5.9) [59] These findings suggest that diabetes may promote a systemic fibrotic milieu, extending beyond internal organs to musculoskeletal tissues. Our findings reinforce this hypothesis, demonstrating that the risk of developing fibrotic hand manifestations is elevated in type 2 diabetes by similar risk as those of internal organs.

We previously hypothesized that the diabetic hand may reflect a systemic profibrotic trajectory-since it is available for clinical examination and could serve as an external clinical biomarker. [60] A recent study employed Mendelian randomization to demonstrate a causal relationship between hyperglycemia duration and all upper limb manifestations. [3] However, whether these manifestations represent distinct conditions, or a single fibrotic process remains uncertain. Conceptualizing them under a unifying term, such as ‘hand fibrosis,’ may help identify individuals at risk for systemic fibrosis. A few studies [9,14,21,28,30–32,40] reported prevalences of all these conditions and allowed us to estimate the ‘overall risk’ of developing any. Despite variations in prevalence, diabetes conferred a similar relative risk across all manifestations. This finding hints at a common underlying pathway rather than independent disease processes.

We used risk ratios (RRs) rather than odds ratios (ORs) to provide a more clinically meaningful assessment of risk. While ORs are often reported in case-control studies, they tend to overestimate risk, particularly when the outcome is not rare. RRs offer a more intuitive measure of disease probability in exposed versus unexposed populations, making them easier to interpret in clinical practice. Since they were calculated using the actual numbers reported, we could homogenize reported metrics across various studies.

Limited joint mobility was first described in type 1 diabetes. [1] Although it is difficult to directly compare across studies, our review shows a higher (1.5 to 2 times) pooled relative risk of developing most manifestations in type 1 diabetes as compared to type 2 diabetes, especially in the earlier studies. Several factors may explain this disparity: Type 1 diabetes typically has an earlier onset and longer disease duration, leading to prolonged exposure to hyperglycemia. Additionally, genetic predisposition may play a role, as type 1 diabetes is associated with human leukocyte antigen (HLA) alleles such as HLA-DR3 and HLA-DR4, which are also linked to autoimmune musculoskeletal conditions. [61] Exogenous insulin, uniformly prescribed in type 1, but not type 2, may contribute to fibrosis through its growth factor-like effects. [62]. Conversely, certain drugs used in type 2 diabetes, such as metformin and sodium-glucose transporter protein 2 (SGLT2) inhibitors, have demonstrated antifibrotic properties, potentially mitigating fibrosis progression. [63] Future meta-regressions incorporating factors such as disease duration, medication use, and genetic background may help elucidate these differences.

The relative risk estimates for hand manifestations in type 2 diabetes were relatively uniform, suggesting that diabetes acts as an amplifying factor rather than an independent trigger. In individuals with an underlying predisposition, diabetes increases the likelihood of developing these conditions. Our data do not determine which individuals are at risk for specific conditions. Potential determinants include discrete genetic risks (such as those identified in genome-wide association studies for Dupuytren’s disease [64] occupational exposures (e.g., repetitive manual labor increasing the risk of CTS, and lifestyle habits (alcohol intake being linked to DD) [65,66] Future meta-regressions examining these factors could help identify subgroups at higher risk for specific manifestations.

Heterogeneity remains a major limitation in the field, as corroborated by earlier meta-analysis for individual hand manifestation [DD 56%, CTS 87%] [5,6]. A major contributory factor to this heterogeneity is likely to be variability in diagnostic criteria and measurement techniques across studies. While this heterogeneity complicates direct comparisons, it also highlights the urgent need for standardized definitions and objective quantification methods in such conditions. We described a potential solution by measuring metacarpophalangeal (MCP) joint extension, independent of specific hand manifestation [67]. Imaging modalities such as magnetic resonance imaging (MRI), as well as artificial intelligence-based quantification methods, could also be explored.

Our analysis shows a decline in pooled risk ratios for diabetic hand manifestations over time, particularly in type 2 diabetes. This trend likely reflects improved glycemic control, earlier detection, and the antifibrotic effects of newer diabetes medications. Increased screening, occupational modifications, and better diabetes management may have further contributed. However, the persistent risk, especially in type 1 diabetes, suggests that longer disease duration and underlying genetic factors play a role. Continued musculoskeletal monitoring and targeted interventions remain essential for reducing long-term complications.

This systematic review has several strengths: It is the first to comprehensively analyze all four major hand manifestations of diabetes within a single framework, allowing for a more integrated understanding of diabetic hand fibrosis. We separately assess risk in both type 1 and type 2 diabetes, providing insights into differential disease susceptibility. The inclusion of a large number of studies strengthens the reliability of our pooled estimates, despite inherent heterogeneity in study design. Furthermore, we standardized the metric of risk reporting by using risk ratios (RRs) instead of odds ratios (ORs), which enhances clinical interpretability. Limitations include the substantial heterogeneity across studies, which reflects differences in diagnostic criteria, study populations, and methodologies. While we attempted to address this by standardizing risk estimates, residual variability remains. Additionally, this review did not account for individual risk factors such as genetic predisposition, duration of diabetes, medication use, occupational exposure, or comorbidities, all of which may influence susceptibility to specific manifestations. Future analyses incorporating meta-regression techniques could help disentangle the contributions of these factors and refine risk stratification for diabetic hand fibrosis. Finally, a meta-analysis of genome-wide association studies (GWAS) across these conditions could help identify shared genetic risk factors, providing insight into common pathways driving fibrosis in diabetes and informing potential therapeutic targets.

In conclusion, our systematic review confirms that diabetes significantly increases the risk of all four major hand manifestations—Dupuytren’s disease, flexor tenosynovitis, limited joint mobility, and carpal tunnel syndrome—by approximately three-fold in type 2 diabetes and up to five-fold in type 1 diabetes. The similar relative risks across manifestations suggest that diabetes uniformly amplifies underlying predispositions rather than acting as a primary trigger. Substantial heterogeneity highlights the need for standardized diagnostic criteria and quantification methods.

## Supporting information

Electronic Supplementary Material

## Acknowledgements

We acknowledge the contribution of Sarita Jadhav and Pranava Nande.

## Data availability

Data shall me made available by the corresponding author upon provision of an analysis plan.

## Funding

DBT/Wellcome India Alliance Fellowship (IA/CPHE/19/1/504607) to Dr Sanat Phatak

## Conflict of interests

None to be declared.

## Contribution statement

All authors significantly contributed to the manuscript and have approved the final version for publication. SH, SP, JLI designed the search strategy, RW, KC, VS, RT, SD, SP contributed to the study selection, data extraction, and qualitative assessment. RW, SK, MK, SP contributed to the data analysis and interpretation of data. SP, KC, RW wrote the initial draft and JLI, CSY, MK critically revised it. SP is the guarantor of this work.

## Notes

### Competing Interest Statement

The authors have declared no competing interest.

