## Supplementary material for "Elevated risk of fibrotic hand manifestations in diabetes: a systematic review and meta-analysis of case-control studies": Electronic Supplementary Material

Electronic Supplementary Data:

INDEX

ESM Table 1: PRISMA (2020) Checklist

| Section and Topic | Item # | Checklist item | Reported (Yes/No) | Location where item is reported |
|---|---|---|---|---|
| **TITLE:** Elevated Risk of fibrotic hand manifestations in diabetes: a systematic review and meta-analysis of case-control studies | | | | |
| Title | 1 | Identify the report as a systematic review. | Yes | Pg 1 |
| **ABSTRACT** | | | | |
| Abstract | 2 | See the PRISMA 2020 for Abstracts checklist. | Yes | Pg 1 |
| **INTRODUCTION** | | | | |
| Rationale | 3 | Describe the rationale for the review in the context of existing knowledge. | Yes | Pg 3 |
| Objectives | 4 | Provide an explicit statement of the objective(s) or question(s) the review addresses. | Yes | Pg 3 |
| **METHODS** | | | | |
| Eligibility criteria | 5 | Specify the inclusion and exclusion criteria for the review and how studies were grouped for the syntheses. | Yes | Pg 4 |
| Informatio n sources | 6 | Specify all databases, registers, websites, organisations, reference lists and other sources searched or consulted to identify studies. Specify the date when each source was last searched or consulted. | Yes | Pg 4 |
| Search strategy | 7 | Present the full search strategies for all databases, registers and websites, including any filters and limits used. | Yes | ESM Table 2 |
| Selection process | 8 | Specify the methods used to decide whether a study met the inclusion criteria of the review, including how many reviewers screened each record and each report retrieved, whether they worked independently, and if applicable, details of automation tools used in the process. | Yes | Pg 4 |
| Data collection process | 9 | Specify the methods used to collect data from reports, including how many reviewers collected data from each report, whether they worked independently, any processes for obtaining or confirming data from study investigators, and if applicable, details of automation tools used in the process. | Yes | Pg 4 |
| Data items | 10a | List and define all outcomes for which data were sought. Specify whether all results that were compatible with each outcome domain in each study were sought (e.g. for all measures, time points, analyses), and if not, the methods used to decide which results to collect. | Yes | Pg 4 |
| | 10b | List and define all other variables for which data were sought (e.g. participant and intervention characteristics, funding sources). Describe any assumptions made about any missing or unclear information. | No | N/A |

| Section and Topic | Item # | Checklist item | Reported (Yes/No) | Location where item is reported |
|---|---|---|---|---|
| Study risk of bias assessment | 11 | Specify the methods used to assess risk of bias in the included studies, including details of the tool(s) used, how many reviewers assessed each study and whether they worked independently, and if applicable, details of automation tools used in the process. | Yes | Pg 5-6 |
| Effect measures | 12 | Specify for each outcome the effect measure(s) (e.g. risk ratio, mean difference) used in the synthesis or presentation of results. | Yes | Pg 5 |
| Synthesis methods | 13a | Describe the processes used to decide which studies were eligible for each synthesis (e.g. tabulating the study intervention characteristics and comparing against the planned groups for each synthesis (item #5)). | Yes | Pg 5 |
| | 13b | Describe any methods required to prepare the data for presentation or synthesis, such as handling of missing summary statistics, or data conversions. | No | N/A |
| | 13c | Describe any methods used to tabulate or visually display results of individual studies and syntheses. | Yes | Pg 6-7 |
| | 13d | Describe any methods used to synthesize results and provide a rationale for the choice(s). If meta-analysis was performed, describe the model(s), method(s) to identify the presence and extent of statistical heterogeneity, and software package(s) used. | Yes | Pg 5 |
| | 13e | Describe any methods used to explore possible causes of heterogeneity among study results (e.g. subgroup analysis, meta-regression). | Yes | Pg 5 |
| | 13f | Describe any sensitivity analyses conducted to assess robustness of the synthesized results. | No | N/A |
| Reporting bias assessment | 14 | Describe any methods used to assess risk of bias due to missing results in a synthesis (arising from reporting biases). | No | N/A |
| Certainty assessment | 15 | Describe any methods used to assess certainty (or confidence) in the body of evidence for an outcome. | No | N/A |
| **RESULTS** | | | | |
| Study selection | 16a | Describe the results of the search and selection process, from the number of records identified in the search to the number of studies included in the review, ideally using a flow diagram. | Yes | ESM Fig 1 |
| | 16b | Cite studies that might appear to meet the inclusion criteria, but which were excluded, and explain why they were excluded. | Yes | ESM Fig 1 |
| Study characteristics | 17 | Cite each included study and present its characteristics. | Yes | Table 1s |
| Risk of bias in studies | 18 | Present assessments of risk of bias for each included study. | Yes | ESM Table 3 |
| Results of individual studies | 19 | For all outcomes, present, for each study: (a) summary statistics for each group (where appropriate) and (b) an effect estimate and its precision (e.g. confidence/credible interval), ideally using structured tables or plots. | Yes | Pg 6-7 |

| Section and Topic | Item # | Checklist item | Reported (Yes/No) | Location where item is reported |
|---|---|---|---|---|
| Results of syntheses | 20a | For each synthesis, briefly summarise the characteristics and risk of bias among contributing studies. | Yes | Pg 7, Table 1, ESM Table 3 |
| | 20b | Present results of all statistical syntheses conducted. If meta-analysis was done, present for each the summary estimate and its precision (e.g. confidence/credible interval) and measures of statistical heterogeneity. If comparing groups, describe the direction of the effect. | Yes | Pg 6-7 |
| | 20c | Present results of all investigations of possible causes of heterogeneity among study results. | No | N/A |
| | 20d | Present results of all sensitivity analyses conducted to assess the robustness of the synthesized results. | No | N/A |
| Reporting biases | 21 | Present assessments of risk of bias due to missing results (arising from reporting biases) for each synthesis assessed. | No | N/A |
| Certainty of evidence | 22 | Present assessments of certainty (or confidence) in the body of evidence for each outcome assessed. | No | N/A |
| **DISCUSSION** | | | | |
| Discussion | 23a | Provide a general interpretation of the results in the context of other evidence. | Yes | Pg 8-11 |
| | 23b | Discuss any limitations of the evidence included in the review. | Yes | Pg 10 |
| | 23c | Discuss any limitations of the review processes used. | Yes | Pg 10 |
| | 23d | Discuss implications of the results for practice, policy, and future research. | Yes | Pg 10-11 |
| **OTHER INFORMATION** | | | | |
| Registration and protocol | 24a | Provide registration information for the review, including register name and registration number, or state that the review was not registered. | Yes | Pg 4 |
| | 24b | Indicate where the review protocol can be accessed, or state that a protocol was not prepared. | Yes | Pg 6 |
| | 24c | Describe and explain any amendments to information provided at registration or in the protocol. | | |
| Support | 25 | Describe sources of financial or non-financial support for the review, and the role of the funders or sponsors in the review. | Yes | Pg 11 |
| Competing interests | 26 | Declare any competing interests of review authors. | Yes | Pg 11 |
| Availability of data, code and other materials | 27 | Report which of the following are publicly available and where they can be found: template data collection forms; data extracted from included studies; data used for all analyses; analytic code; any other materials used in the review. | No | N/A |

*From:* Page MJ, McKenzie JE, Bossuyt PM, Boutron I, Hoffmann TC, Mulrow CD, et al. The PRISMA 2020 statement: an updated guideline for reporting systematic reviews. BMJ 2021;372:n71. doi: 10.1136/bmj.n71. This work is licensed under CC BY 4.0. To view a copy of this license, visit https://creativecommons.org/licenses/by/4.0/

PRISMA 2020 Abstracts Checklist:

| Section and Topic | Item # | Checklist item | Reported (Yes/No) |
|---|---|---|---|
| **TITLE** | | | |
| Title | 1 | Identify the report as a systematic review. | Yes |
| **BACKGROUND** | | | |
| Objectives | 2 | Provide an explicit statement of the main objective(s) or question(s) the review addresses. | Yes |
| **METHODS** | | | |
| Eligibility criteria | 3 | Specify the inclusion and exclusion criteria for the review. | Yes |
| Information sources | 4 | Specify the information sources (e.g. databases, registers) used to identify studies and the date when each was last searched. | No |
| Risk of bias | 5 | Specify the methods used to assess risk of bias in the included studies. | No |
| Synthesis of results | 6 | Specify the methods used to present and synthesise results. | No |
| **RESULTS** | | | |
| Included studies | 7 | Give the total number of included studies and participants and summarise relevant characteristics of studies. | Yes |
| Synthesis of results | 8 | Present results for main outcomes, preferably indicating the number of included studies and participants for each. If meta-analysis was done, report the summary estimate and confidence/credible interval. If comparing groups, indicate the direction of the effect (i.e. which group is favoured). | Yes |
| **DISCUSSION** | | | |
| Limitations of evidence | 9 | Provide a brief summary of the limitations of the evidence included in the review (e.g. study risk of bias, inconsistency and imprecision). | Yes |
| Interpretation | 10 | Provide a general interpretation of the results and important implications. | Yes |
| **OTHER** | | | |
| Funding | 11 | Specify the primary source of funding for the review. | Yes |
| Registration | 12 | Provide the register name and registration number. | Yes |

*From:* Page MJ, McKenzie JE, Bossuyt PM, Boutron I, Hoffmann TC, Mulrow CD, et al. The PRISMA 2020 statement: an updated guideline for reporting systematic reviews. BMJ 2021;372:n71. doi: 10.1136/bmj.n71. This work is licensed under CC BY 4.0. To view a copy of this license, visit https://creativecommons.org/licenses/by/4.0/

ESM Table 2: Search strategy details

| Database: Medline (PubMed) | | |
|---|---|---|
| 1. | "Diabetes Mellitus"[Mesh] OR "Diabetes Complications"[Mesh] OR "Prediabetic State"[Mesh] OR "Glucose Intolerance"[Mesh] OR "Insulin Resistance"[Mesh] OR diabetes[tiab] OR diabetic[tiab] OR prediabetes[tiab] OR prediabetic[tiab] OR "insulin resistance"[tiab] OR "insulin insensitivity"[tiab] OR "glucose intolerance"[tiab] OR "impaired glucose intolerance"[tiab] | 806787 |
| 2. | "Carpal Tunnel Syndrome"[Mesh] OR "Dupuytren Contracture"[Mesh] OR "trigger finger"[tiab] OR "trigger fingers"[tiab] OR "carpal tunnel"[tiab] OR LJM[tiab] OR "limited joint mobility"[tiab] | 15787 |
| 3. | "Hand"[Mesh] OR "Hand Joints"[Mesh] OR "Hand Bones"[Mesh] OR hand[tiab] OR hands[tiab] OR fingers[tiab] OR finger[tiab] OR dupuytren[tiab] OR dupuytren's[tiab] OR "median nerve"[tiab] OR "median nerves"[tiab] OR flexor[tiab] | 588131 |
| 4. | "Fibrosis"[Mesh] OR "Contracture"[Mesh] OR "Tenosynovitis"[Mesh] OR fibrosis[tiab] OR contracture[tiab] OR contractures[tiab] OR syndrome[tiab] OR syndrome[tiab] OR tenosynovitis[tiab] OR tenovaginosis OR fibrosis[tiab] OR cheiroarthropathy[tiab] OR cheiroarthropathies[tiab] | 1314038 |
| 5. | 2 OR (3 AND 4) | 54192 |
| 6. | 1 AND 5 | 2818 |

| | | |
|---|---|---|
| **Database: Embase (Elsevier)** | | |
| 1. | 'diabetes mellitus'/exp OR 'impaired glucose tolerance'/exp OR 'glucose intolerance'/exp OR 'insulin resistance'/exp OR (diabetes OR diabetic OR prediabetes OR prediabetic OR 'insulin resistance' OR 'insulin insensitivity' OR 'glucose intolerance'):ti,ab | 1320845 |
| 2. | 'carpal tunnel syndrome'/exp OR 'Dupuytren contracture'/exp OR 'trigger finger'/exp OR ('trigger finger' OR 'trigger fingers' OR 'carpal tunnel' OR LJM OR 'limited joint mobility'):ti,ab | 32030 |
| 3. | 'hand'/exp OR 'hand joint'/exp OR 'hand bone'/exp OR (hand OR hands OR fingers OR finger OR dupuytren OR dupuytrens OR 'median nerve' OR 'median nerves' OR flexor):ti,ab | 761850 |
| 4. | 'fibrosis'/exp OR 'contracture'/exp OR 'tenosynovitis'/exp OR 'fibrosis'/exp OR (fibrosis OR contracture OR contractures OR syndrome OR syndrome OR tenosynovitis OR tenovaginosis OR fibrosis OR cheiroarthropathy OR cheiroarthropathies):ti,ab | 1746066 |
| 5. | 2 OR (3 AND 4) | 79348 |
| 6. | 1 AND 5 | 5459 |
| **Database: CINAHL (EbscoHost)** | | |

| | | |
|---|---|---|
| 1. | MH "Diabetes Mellitus+" OR MH "Prediabetic State" OR MH "Glucose Intolerance" OR MH "Insulin Resistance+" OR TI(diabetes OR diabetic OR prediabetes OR prediabetic OR "insulin resistance" OR "insulin insensitivity" OR "glucose intolerance" OR "impaired glucose intolerance") OR AB(diabetes OR diabetic OR prediabetes OR prediabetic OR "insulin resistance" OR "insulin insensitivity" OR "glucose intolerance" OR "impaired glucose intolerance") | 268280 |
| 2. | MH "Carpal Tunnel Syndrome" OR MH "Dupuytren's Contracture" OR TI("trigger finger" OR "trigger fingers" OR "carpal tunnel" OR LJM OR "limited joint mobility") OR AB("trigger finger" OR "trigger fingers" OR "carpal tunnel" OR LJM OR "limited joint mobility") | 4547 |
| 3. | MH "Hand+" OR MH "Hand Joints+" OR TI(hand OR hands OR fingers OR finger OR dupuytren OR dupuytren's OR "median nerve" OR "median nerves" OR flexor) OR AB(hand OR hands OR fingers OR finger OR dupuytren OR dupuytren's OR "median nerve" OR "median nerves" OR flexor) | 102098 |
| 4. | MH "Fibrosis" OR MH "Contracture+" OR MH "Tenosynovitis+" OR TI(fibrosis OR contracture OR contractures OR syndrome OR syndrome OR tenosynovitis OR tenovaginosis OR fibrosis OR cheiroarthropathy OR cheiroarthropathies) OR AB(fibrosis OR contracture OR contractures OR syndrome OR syndrome OR tenosynovitis OR tenovaginosis OR fibrosis OR cheiroarthropathy OR cheiroarthropathies) | 242391 |
| 5. | 2 OR (3 AND 4) | 10686 |
| 6. | 1 AND 5 | 623 |
| **Database: Scopus (Elsevier)** | | |

| | | |
|---|---|---|
| 1. | TITLE-ABS(diabetes OR diabetic OR prediabetes OR prediabetic OR {insulin resistance} OR {insulin insensitivity} OR {glucose intolerance} OR {impaired glucose intolerance}) | 867542 |
| 2. | TITLE-ABS({trigger finger} OR {trigger fingers} OR {carpal tunnel} OR LJM OR {limited joint mobility}) | 13693 |
| 3. | TITLE-ABS(hand OR hands OR fingers OR finger OR dupuytren OR dupuytren's OR {median nerve} OR {median nerves} OR flexor) | 1384198 |
| 4. | TITLE-ABS(fibrosis OR contracture OR contractures OR syndrome OR syndrome OR tenosynovitis OR tenovaginosis OR fibrosis OR cheiroarthropathy OR cheiroarthropathies) | 1456916 |
| 5. | 2 OR (3 AND 4) | 55892 |
| 6. | 1 AND 5 | 2814 |
| **Database: Clinicaltrials.gov** | | |
| 1. | (diabetes OR diabetic OR prediabetes OR prediabetic OR "insulin resistance" OR "insulin insensitivity" OR "glucose intolerance" OR "impaired glucose intolerance") | 19906 |
| 2. | "trigger finger" OR "trigger fingers" OR "carpal tunnel" OR LJM OR "limited joint mobility" OR hand OR hands OR fingers OR finger OR dupuytren OR dupuytren's OR "median nerve" OR "median nerves" | 1981 |
| 3. | 1 AND 2 | 628 |

ESM Table 3: Risk of Bias Assessment Using the Newcastle-Ottawa Scale

| Author (Publication year) | Selection | Definition of Cases | Representativeness of cases | Selection of Controls | Definition of Controls | Comparability | Comparability of Cases & Controls on basis of design/ analysis | Exposure and outcomes | Ascertainment of exposure | Same method of ascertainment for cases and controls | Non-Response rate | NOS Score |
|---|---|---|---|---|---|---|---|---|---|---|---|---|
| **Type 2 diabetes** | | | | | | | | | | | | |
| Akyol et al 2006 | *** | 1 | 1 | 0 | 1 | * | 1 | ** | 1 | 1 | 0 | 6 |
| Abogamal et al 2017 | ** | 1 | 0 | 0 | 1 | * | 1 | ** | 1 | 1 | 0 | 5 |
| Ardic et al 2003 | ** | 1 | 0 | 0 | 1 | * | 1 | * | 0 | 1 | 0 | 4 |
| Aydeniz et al 2008 | *** | 1 | 1 | 0 | 1 | * | 1 | ** | 1 | 1 | 0 | 6 |
| Cagliero et al 2002 | *** | 1 | 1 | 0 | 1 | * | 1 | *** | 1 | 1 | 1 | 7 |
| Cederlund et al 2009 | *** | 1 | 0 | 1 | 1 | * | 1 | ** | 1 | 1 | 0 | 6 |
| Chammas et al 1995 | *** | 1 | 0 | 1 | 1 | * | 1 | * | 0 | 1 | 0 | 5 |
| Eadington et al 1991 | * | 0 | 0 | 0 | 1 | * | 1 | ** | 1 | 1 | 0 | 6 |
| Fernando et al 1997 | *** | 0 | 1 | 1 | 1 | * | 1 | * | 0 | 1 | 0 | 5 |
| Fitzcharles et al 1984 | ** | 0 | 0 | 1 | 1 | * | 1 | * | 0 | 1 | 0 | 4 |
| Gertner et al 1990 | * | 0 | 0 | 0 | 1 | * | 1 | ** | 1 | 1 | 0 | 4 |
| Ikem et al 2009 | * | 0 | 0 | 0 | 1 | * | 1 | * | 0 | 1 | 0 | 3 |
| Kermani et al 2017 | ** | 1 | 0 | 0 | 1 | * | 1 | ** | 1 | 1 | 0 | 6 |
| Kidwai et al 2013 | ** | 1 | 1 | 0 | 0 | ** | 2 | ** | 1 | 1 | 0 | 6 |
| Kovacs et al 2012 | ** | 0 | 0 | 1 | 1 | * | 1 | * | 0 | 1 | 0 | 4 |
| Larkin et al 1986 | ** | 0 | 1 | 0 | 1 | * | 1 | ** | 1 | 1 | 0 | 5 |
| Lu et al 1993 | ** | 1 | 0 | 0 | 1 | * | 1 | ** | 1 | 1 | 0 | 5 |
| Mengist et al 1985 | ** | 0 | 1 | 0 | 1 | * | 1 | ** | 1 | 1 | 0 | 5 |
| Naha et al 2021 | ** | 0 | 0 | 1 | 1 | * | 1 | ** | 1 | 1 | 0 | 5 |
| Oktayoglu et al 2015 | ** | 1 | 0 | 0 | 1 | * | 1 | ** | 1 | 1 | 0 | 5 |

| | | | | | | | | | | | |
|---|---|---|---|---|---|---|---|---|---|---|---|
| Olaosebikan 2020 | ** | 0 | 0 | 1 | 1 | * | 1 | * | 0 | 1 | 0 | 4 |
| Pal et al 1986 | *** | 1 | 1 | 0 | 1 | * | 1 | ** | 1 | 1 | 0 | 6 |
| Pandey et al 2013 | ** | 1 | 0 | 0 | 1 | * | 1 | ** | 1 | 1 | 0 | 5 |
| Ravindran Rajendran et al 2011 | ** | 0 | 0 | 1 | 1 | * | 1 | * | 0 | 1 | 0 | 4 |
| Renard et al 1994 | ** | 1 | 0 | 0 | 1 | * | 1 | ** | 1 | 1 | 0 | 5 |
| Saini et al 2016 | *** | 1 | 1 | 0 | 1 | * | 1 | ** | 1 | 1 | 0 | 6 |
| Savaş et al 2007 | * | 0 | 0 | 0 | 1 | * | 1 | * | 0 | 1 | 0 | 3 |
| Starkman et al 1986 | * | 0 | 0 | 0 | 1 | * | 1 | ** | 1 | 1 | 0 | 4 |
| **Type 1 diabetes** | | | | | | | | | | | | |
| Arkkila et al *1994 | *** | 0 | 1 | 1 | 1 | * | 1 | ** | 1 | 1 | 0 | 6 |
| Cagliero et al 2002 | *** | 1 | 1 | 0 | 1 | * | 1 | *** | 1 | 1 | 1 | 7 |
| Chammas et al 1995 | *** | 1 | 0 | 1 | 1 | * | 1 | * | 0 | 1 | 0 | 5 |
| Clarke et al 1990 | **** | 1 | 1 | 1 | 1 | * | 1 | ** | 1 | 1 | 0 | 7 |
| Eadington et al 1989 | * | 0 | 0 | 0 | 1 | * | 1 | ** | 1 | 1 | 0 | 4 |
| Garza-Elizondo et al 1983 | ** | 0 | 0 | 1 | 1 | ** | 2 | ** | 1 | 1 | 0 | 6 |
| Gertner et al 1990 | * | 0 | 0 | 0 | 1 | * | 1 | ** | 1 | 1 | 0 | 4 |
| Gutefeldt et al 2019 | ** | 0 | 0 | 1 | 1 | * | 1 | * | 0 | 1 | 0 | 4 |
| Haitas et al 1986 | * | 0 | 0 | 0 | 1 | * | 1 | * | 0 | 1 | 0 | 3 |
| Holte et al 2017 | ** | 0 | 0 | 1 | 1 | * | 1 | * | 0 | 1 | 0 | 4 |
| Kakourou et al 1994 | *** | 1 | 0 | 1 | 1 | * | 1 | ** | 1 | 1 | 0 | 6 |
| Kennedy et al 1982 | ** | 1 | 0 | 0 | 1 | * | 1 | ** | 1 | 1 | 0 | 5 |
| Kovacs et al 2012 | ** | 0 | 0 | 1 | 1 | * | 1 | * | 0 | 1 | 0 | 4 |
| Labad et al 2018 | ** | 0 | 0 | 1 | 1 | * | 1 | ** | 0 | 1 | 1 | 5 |
| Mengistu et al 1985 | ** | 0 | 1 | 0 | 1 | * | 1 | ** | 1 | 1 | 0 | 5 |
| Montana et al 1995 | * | 0 | 0 | 0 | 1 | * | 1 | ** | 1 | 1 | 0 | 4 |
| Pal et al 1986 | *** | 1 | 1 | 0 | 1 | * | 1 | ** | 1 | 1 | 0 | 6 |
| Raje et al 2015 | ** | 0 | 0 | 1 | 1 | * | 1 | * | 0 | 1 | 0 | 4 |
| Renard 1994 | ** | 1 | 0 | 0 | 1 | * | 1 | ** | 1 | 1 | 0 | 5 |
| Sauseng et al 2002 | ** | 1 | 0 | 0 | 1 | * | 1 | * | 0 | 1 | 0 | 4 |
| Vukovic et al 1996 | *** | 1 | 0 | 1 | 1 | * | 1 | * | 0 | 1 | 0 | 5 |

| | | | | | | | | | | | |
|---|---|---|---|---|---|---|---|---|---|---|---|
| Yosipovitch et al 1990 | * | 0 | 0 | 0 | 1 | * | 1 | ** | 1 | 1 | 0 | 4 |
| Starkman et al 1986 | * | 0 | 0 | 0 | 1 | * | 1 | ** | 1 | 1 | 0 | 4 |
| **Diabetes (type unspecified)*** | | | | | | | | | | | | |
| Ravid et al 1977 | ** | 1 | 0 | 0 | 1 | * | 1 | ** | 1 | 1 | 0 | 5 |
| Brice et al 1982 | ** | 0 | 0 | 1 | 1 | * | 1 | * | 0 | 1 | 0 | 4 |
| Leden et al 1983 | *** | 0 | 1 | 1 | 1 | * | 1 | * | 0 | 1 | 0 | 5 |
| Pal et al 1987 | * | 0 | 0 | 0 | 1 | * | 1 | * | 0 | 1 | 0 | 3 |
| Grandizio et al 2014 | ** | 0 | 0 | 1 | 1 | * | 1 | ** | 1 | 1 | 0 | 5 |
| Font et al 2014 | ** | 0 | 0 | 1 | 1 | * | 1 | * | 0 | 1 | 0 | 4 |
| Lee et al 2018 | ** | 1 | 0 | 0 | 1 | * | 1 | ** | 1 | 1 | 0 | 5 |

ESM Figure 1: PRISMA flow diagram for study selection process

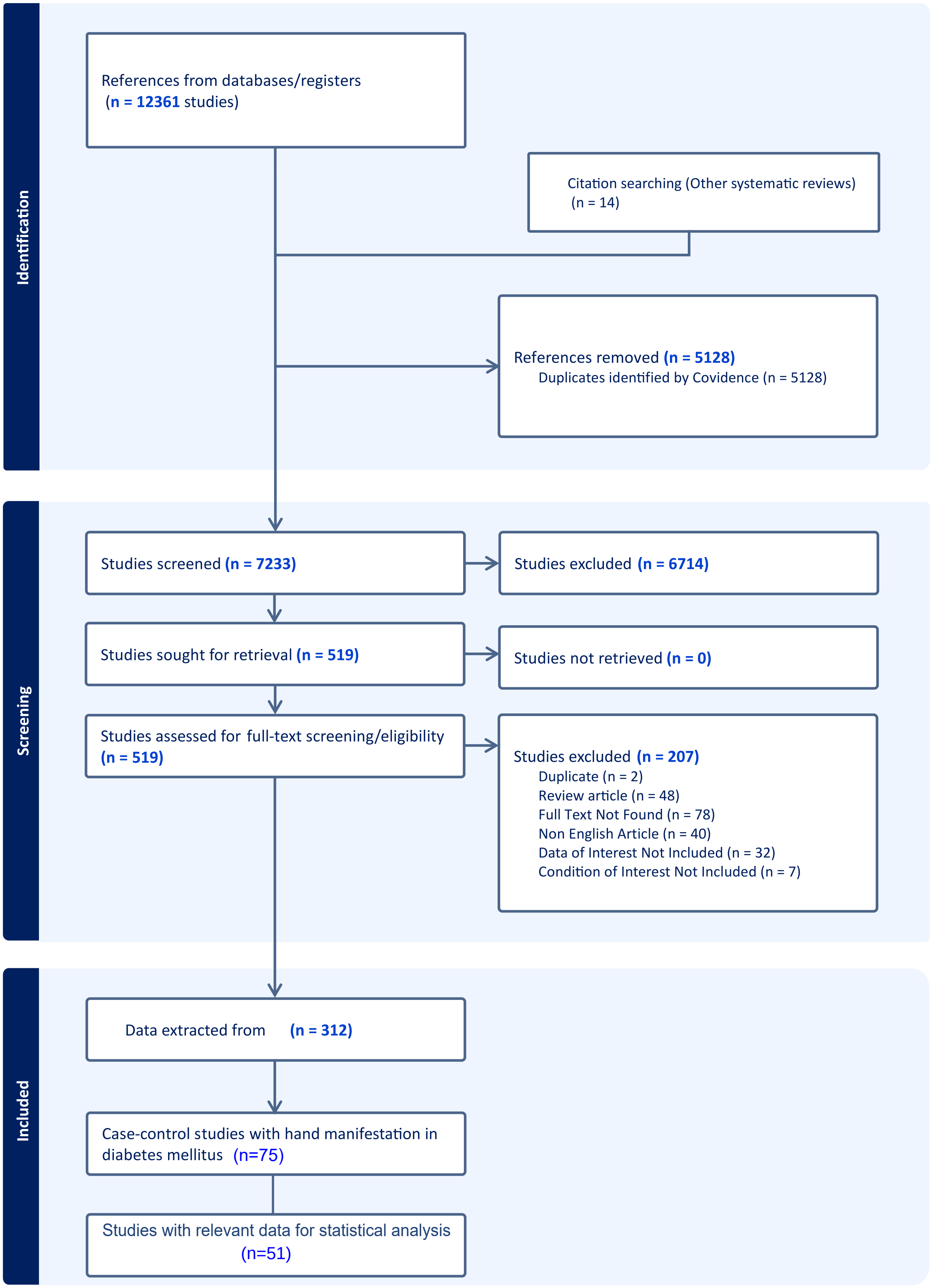
